# An *in silico* integrative analysis of hepatocellular carcinoma omics databases shows the dual regulatory role of microRNAs either as tumor suppressors or as oncomirs

**DOI:** 10.1101/2021.04.27.21256224

**Authors:** Mahmoud M. Tolba, Bangli Soliman, Abdul Jabbar, HebaT’Allah Nasser, Mahmoud Elhefnaw

## Abstract

MicroRNAs are well known as short RNAs bases, 22 nucleotides, binding directly to 3’untranslated region (3’UTR) of the messenger RNA to repress their functions. Recently, microRNAs have been widely used as a therapeutic approach for various types of Cancer. MicroRNA is categorized into tumor suppressor and oncomirs. Tumor suppressor microRNAs can repress the pathologically causative oncogenes of the hallmarks of Cancer. However, based on the fact that miRNA has no proper fidelity to bind specific mRNAs due to binding to off-targets, it results in a kind of inverse biological activity. Here, we have executed an *in-silico* integrative analysis of GEO/TCGA-LIHC of genes/microRNAs expression analysis in HCC, including (446 HCC vs. 146 normal specimens for miRNAs expression) ad 488 specimens for genes expression. It virtually shows that microRNAs could have an ability to target both oncogenes and tumor suppressor genes that contribute to its dual activity role as a tumor suppressor and oncomirs via miRNA-lncRNA-TFs-PPI Crosstalk. Seven resultant microRNAs show a putative dual role in HCC. It enhances our concluded suggestion of using combination therapy of tumor suppressor genes activators with microRNAs.

## 1. Introduction

Different types of Cancer are the lead diseases where the scientific community hunts for new approaches to cure them. One of these approaches is epigenetic therapy [1]. Epigenetics is a genetic field that strengthens our knowledge to comprehensively understand the factors that control the genes translations within the cells [2]. Long non-coding RNAs, small interfering RNA, and microRNAs are among these factors that control the translation of the genes [3-5]. This activity controls various biological processes such as cellular differentiation, cell signaling, life span, and apoptosis. Cancer is a disease of deregulation of the normal state of biological processes called cancer hallmarks [6].

Here we are focusing on Hepatocellular carcinoma (HCC), which is promptly considered one of the most prevalent cancers worldwide [7-9]. With a rising rate, it is a prominent source of mortality. Patients with advanced fibrosis, predominantly cirrhosis, and hepatitis B are predisposed to developing HCC. Individuals with chronic hepatitis B and C infections are most commonly suffering [10]. Different therapeutic options, including liver resection, transplantation, systemic and local therapy, must be tailored to each patient [11]. One of the most promising therapies for HCC is microRNAs therapy. It is well demonstrated that tens of microRNAs have an ectopic expression in HCC. It directly correlates with the cancerous pathway by inducing the list of oncogenes list with the cancerous cells. The therapeutic approach by the restoration of microRNAs is inclined to suppress the oncogenes, decreasing Cancer [12].

### 1.1 miRNAs

MicroRNA dates back to the year 1993 as the first time the microRNAs term has been enlightening. The first microRNA ever was c.elegan let 7. It regulates the c.elegan timing development [13].

MicroRNAs are classes of non-coding RNAs that play an important role in gene expression regulation in Cancer [14, 15]. They are generally transcribed from DNA sequences into primary miRNA and processed into precursor miRNAs, and finally mature miRNAs [3]. The principal role of microRNAs is to control protein translation by binding to complementary sequences in the 3′ untranslated region (UTR) of target messenger RNAs (mRNAs) and thus deleteriously regulate mRNA translation. The microRNA–mRNA binding site is short (6–8 base pairs), and then each microRNA has the prospective to target multiple different mRNAs [16, 17].

### 1.2 MicroRNA Biogenesis

MicroRNAs biosynthesis has been well studied. The more well-known biogenesis pathway is the canonical pathway by which microRNAs are processed [18]. The canonical biogenesis pathway can be described as follows: pri-microRNAs are transcripted from their genes. This process into pri-miRNAs by microprocessor complex (RNA Binding protein DiGeorge Syndrome Critical Region 8 (DGCR8) and a ribonuclease III enzyme, Drosha. DGCR8 recognizes an N6-methyladenylated GGAC and other motifs within the pri-miRNA, while Drosha cleaves the pri-miRNA duplex at the base of the characteristic hairpin structure of pri-miRNA. Accordingly, two nt 3′ overhang on pre-miRNA is formed, one pre-miRNAs are formed. They will be exported by exportin 5 (XPO5)/RanGTP complex and then processed by the RNase III endonuclease Dicer. This processing includes terminal loop removal, and consequently, immature miRNAs duplex is formed.

miRNA strand directionality determines the name of the mature form of miRNA.The 5p strand arises from the 5-end from pre-miRNA hairpin, while the 3p strand originates from the 3-end. The strands mentioned above were derived from the mature miRNA duplex, which can be loaded into the Argonaute (AGO) family of proteins (AGO1-4 in humans) in an ATP-dependent manner. For any given miRNA, this is a great difference between the proportional of AGO-loaded 5p or 3p, which depends on the cell type or cellular environment. They may occur as one more predominant than the other. The 5p or 3p strand selection is based on the thermodynamic stability at the 5′ ends of the miRNA duplex or a 5′ U at nucleotide position1. Commonly, the lower 5-strand or 5-uracil strand is likely loaded into AGO and estimated the guide strand. The unloaded strand (passenger strand) is the unwanted strand from the guide strand through various mechanisms depending on the complementarity degree. The microRNA passenger strand with no mismatches is AGO2 sliced and cellular machinery degraded. Therefore, strong bias can be produced. On the other hand, central mismatches miRNA duplexes or non-AGO2 loaded miRNA are inertly unwound and degraded [19-21].

### 1.3 Bioinformatics role in cancer therapy

Through Bioinformatics analysis of microarray and high through genomic data, hundreds of ectopic expression of genes and microRNAs within various cancer types have been discovered via free online Bioinformatics tools that play an effective role in predicting new targets for microRNA and its genetic enrichment [22]. There are many free online tools [23] that show the binding mode of tens of predicted genes for each microRNA, such as RNA22 [24], DIANA [25], and miRanda [26]. It helps most in discovering new targets for the miRNAs for wet-lab experiments. Here, multiple genes/microRNAs expression datasets have been used to uncover the interaction of miRNAs with mRNAs, long non-coding RNA (LncRNA) [27], transcription factors (TFs) [28], and protein-protein interaction (PPI) [29]. For miRNA-mRNA-LncRNA-TFs-PPI crosstalk analysis, the followed work pipeline has helped most in putatively visualize all the factors that may have a control on miRNAs path to be biologically active as oncomiR or tumor suppressor as shown in the following flowchart (Figure 1).

**Figure 1:**
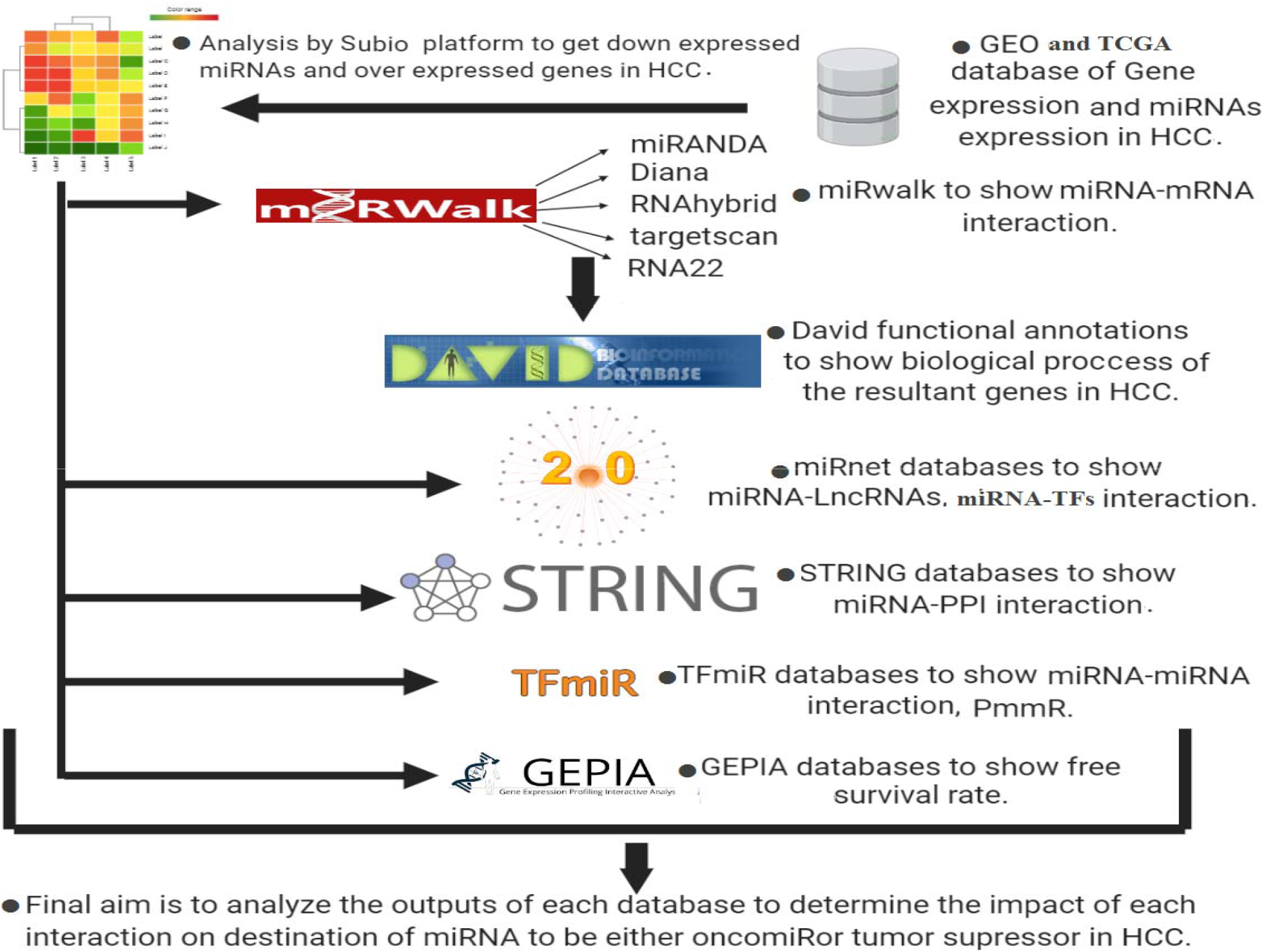
Flowchart of the implemented workflow illustrates the methods that have been followed.

A pool of the most differentially expressed miRs in HCC has been evolved and showed for their involved actions in tumor development or progression. Interestingly in this context, our current study will aim to perform a bioinformatics protocol on a huge list of the significantly down expressed microRNAs within the HCC cells to unveil their activity role. It was found that one microRNA has a potential role in working as a tumor suppressor and tumor activator for the hallmarks of liver cancer.

## 2. Methodology

### 2.1 Collecting data of the significantly down-expressed microRNAs in HCC

Using NCBI Pubmed https://www.ncbi.nlm.nih.gov/pubmed/, we have searched for all published scientific articles and reviews working on microRNAs as a therapeutic approach for liver cancer. We used “MicroRNA+Hepatocellular Carcinoma” as a query for our search. To deepen our data, we utilized the PhenomiR [30] database to get all the microRNAs related to Hepatocellular carcinoma.

To enrich our work, we have surveyed different registry of NCBI Gene Expression Omnibus microarray databases for the miRs, and interestingly we have chosen five databases [31, 32] and one TCGA LIHV –RNA seq databases with broad samples of 446 HCC vs. 146 adjacent normal specimens which cover cirrhosis and G2/M phase of HCC.

These six integrated datasets are “Miltenyi Biotec miRXplore miRNA Microarray, GSE28854’’, Capital Bio Mammalian miRNA Array Services V1 [1].0, GSE10694’’, “Affymetrix Multispecies miRNA-2 Array, GSE74618”Affymetrix Multispecies miRNA-4 Array, GSE115016” “Agilent-031181 Unrestricted_Human_miRNA_V16.0_Microarray, GSE110217” and “TCGA-LIHC, RNA seq’’. These integrated datasets have been downloaded and analyzed by the Subio Platform PC program http://www.subio.jp/products/platform to be filtered to get the overlap of common down-expressed microRNAs in HCC specimens. The resultant list of the down-expressed miRNAs, from datasets analysis, has been cross–confirmed by the previously mentioned manual search within NCBI Pubmed and PhenomiR [33].

### 2.2 Collecting data of over-expressed genes in HCC

We have downloaded three integrated and recent datasets like miR datasets collection by utilizing NCBI GEO microarray databases. They included the following ‘‘ Affymetrix Human Genome U133A 2.0 Array”’ GSE14520”, “Affymetrix HT Human Genome U133A Array, GSE14520” and TCGA-LIHC, and they have been conducted by subio platform program to get the overexpressed genes in HCC.

### 2.3 miRNAs-targets prediction

First, an online miRBase database [34, 35] was used to get access to microRNAs. After that, we have utilized miRWalk [36] to get the predicted and validated target genes of each microRNA. Five different bioinformatics tools have been chosen to get the highest accuracy and specificity for our predicted genes data. These five tools were miRANDA [26], DIANA [25], RNAhybrid [37], targetscan [38] and RNA22 [24]. The validated miRNA targets by miRWalk have been further enriched and confirmed via online experimental miRs-targets interaction databases such as DIANA-TarBase v7.0 [39] and mirtarbase databases [40].

### 2.4 Enrichment and gene ontology

David functional annotation [41] was run to get biological processes and genes ontology of each list of miRNA targets, which resulted from miRwalk. P-value < 0.05 was set as cut off of the chosen biological process. The common resultant genes from datasets analysis by subio platform and shown by miRwalk are enriched by David functional annotation.

The final step was data-filtration to highlight the biological process that is only related to hepatocellular carcinoma hallmarks to show the impact of microRNAs as either tumor suppressor or oncomiR. The previously filtrated genes have been used to draw a microRNAs-targets network by Cytoscape [42] to show the transcriptomic level’s miRs interactions.

### 2.5 miRNA-lncRNA interaction

miRNet online tool [43] was used to determine the regulatory interaction of microRNA-LncRNA and microRNA-TFs interaction.

### 2.6 miRNA-miRNA interaction

TFmiR online tool [44] was used to get the regulatory roles of microRNA-microRNA interaction. TFmiR is based on disease-specific microRNAs of putative miRNA–miRNA regulations, PmmR database [45].

### 2.7 miRNA-‘‘Protein-Protein’’ interaction

Differential gene expression (UP/down) has been used for PPI constructing network by STRING v10 tool [46]. The Biological processes of the resultant network were filtrated into the ones related to HCC. The genes incorporated in the output, Biological processes, were reanalyzed via miRWalk to get down expressed miRNAs in HCC, correlated to these BPs.

### 2.8 Free survival analysis

Here, the aim is to visualize the potential impact of microRNA on the HCC survival progression using GEPIA [47]. The selected genes on the central role of PPI by owning the highest number of edges.

## 3. Results

### 3.1. MicroRNAs ectopic expression in HCC

A list of 24 significantly down expressed microRNAs in HCC has been identified by Subio platform PC for the overall six datasets, including GEO and TCGA-LIHC, RNA seq database (Figure 2). These down expressed microRNAs have been selected because they are common among at least two datasets out of the six datasets. They have been further confirmed via the PhenomiR database (HCC).

**Figure2:**
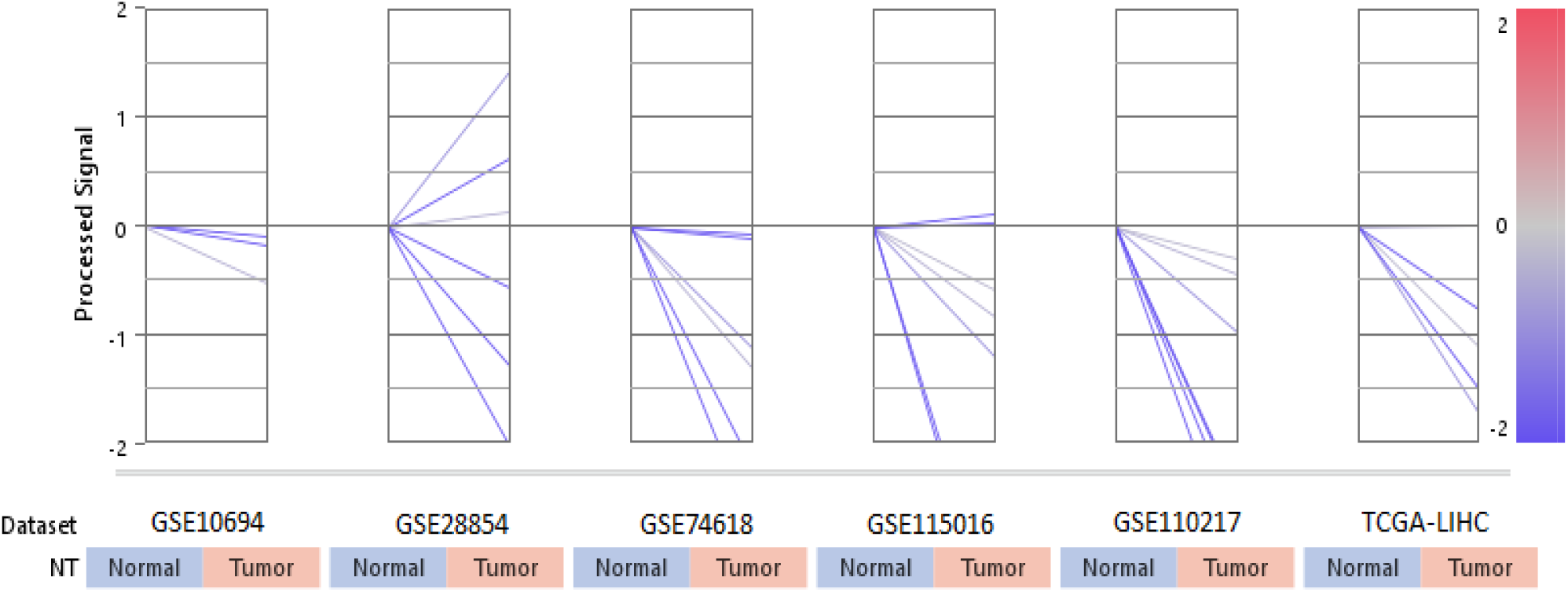
the down-expressed microRNAs in HCC, 24 microRNAs.

### 3.2. Expression of genes targeted by the microRNAs

As one of the main criteria for microRNA is that it is used as a gene therapy, the targeted genes should be overexpressed within the HCC cell line. The overexpression of these genes alters its function to be cancerous genes via inducing cancerous biological processes. Here, GEO datasets and TCGA-LIHC database have been analyzed by subio platform to show the common up-expressed genes in all datasets, combined to HCC target genes which have previously shown by Disease-annotation of miRwalk for Hepatocellular carcinoma. It resulted in 180 overexpressed shared within,’’ Affymetrix Human Genome U133A 2.0 Array” ‘ GSE14520”, “Affymetrix HT Human Genome U133A Array, GSE14520” and TCGA-LIHC (Figure 3 a & b). Any gene, shown as down-expressed in HCC, has been excluded.

**Figure 3:**
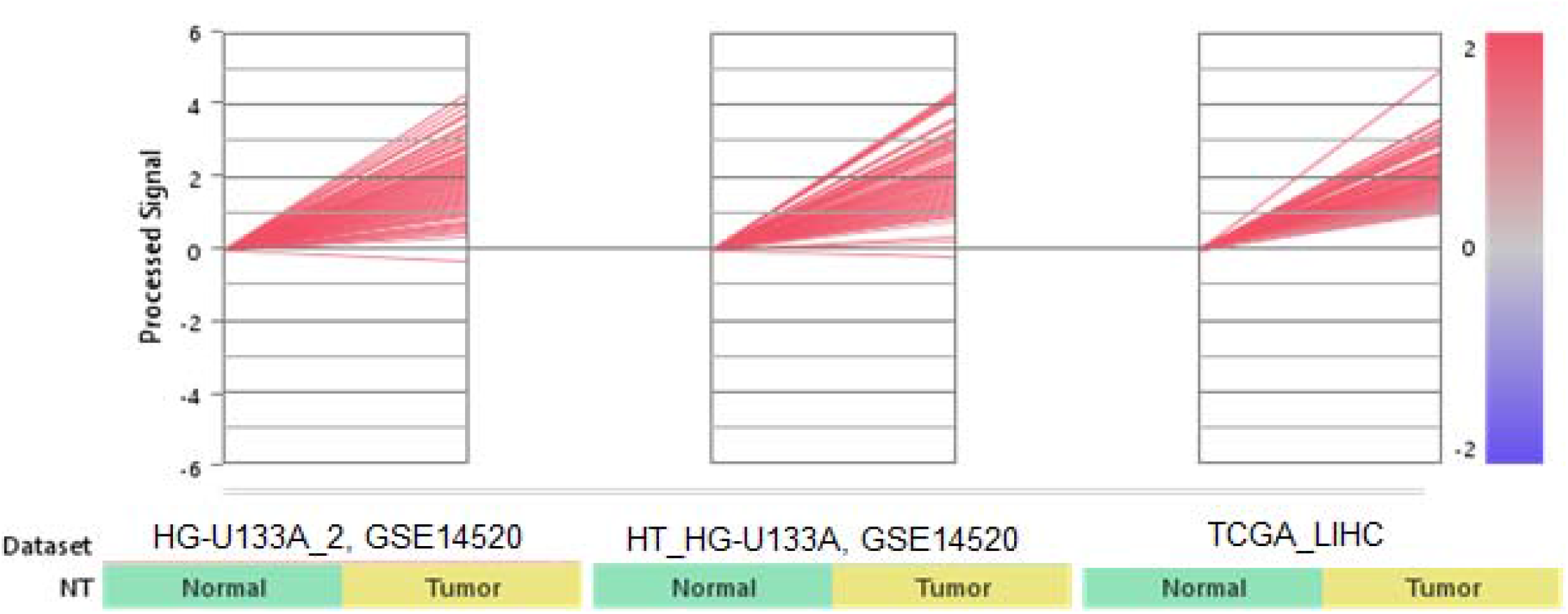
a) the differentially up-expressed genes in HCC

**Figure 3:**
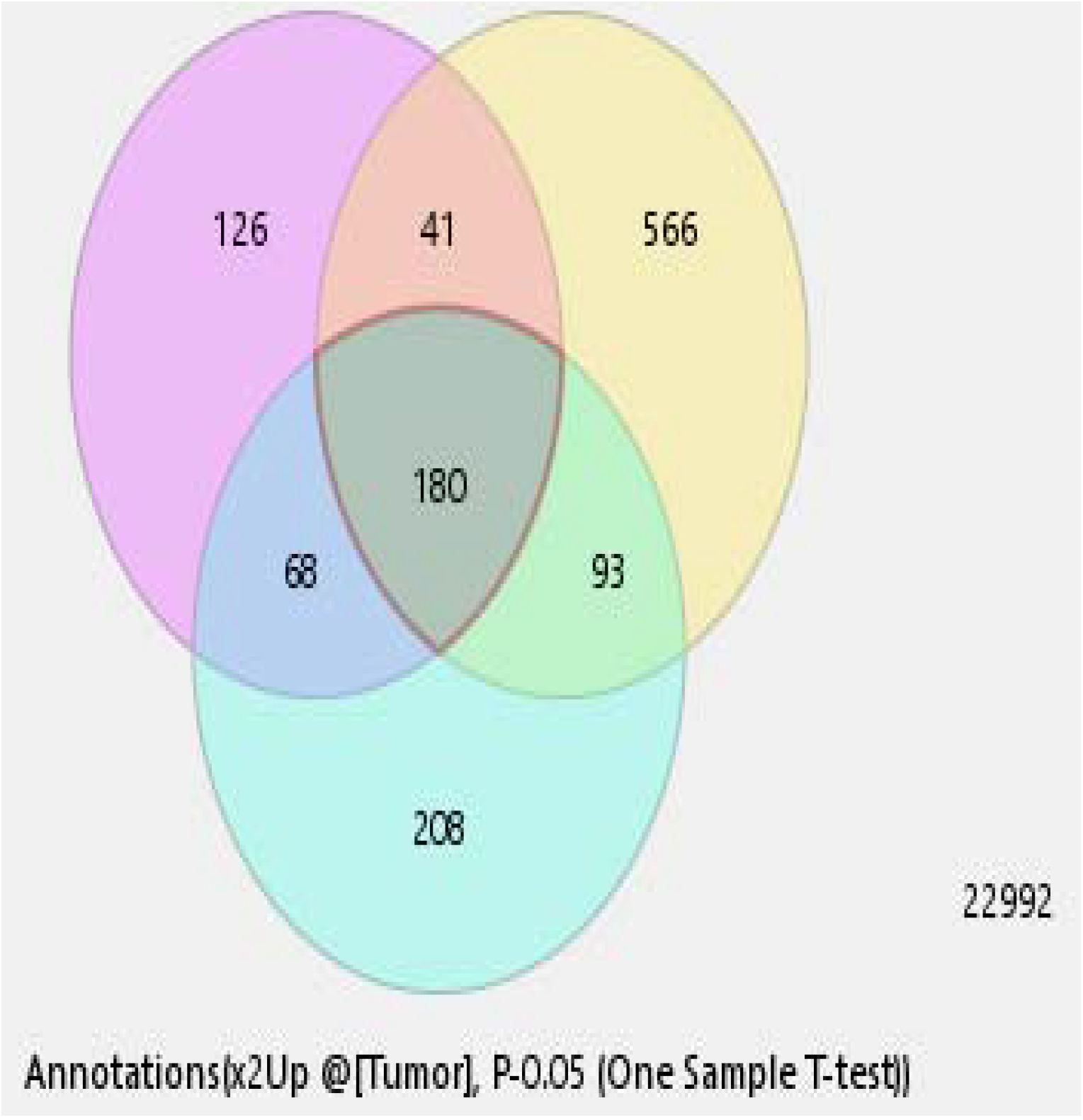
b) 180 genes are found to be common over-expression genes in HCC.

### 3.3. microRNAs-genes interactions

Each microRNA of our 24 ones has hundreds of target genes resulted from an integrative analysis of HCC gene expression datasets by subioplatform (Figure 2), combined with resultant genes from miRwalk related to HCC hallmarks, Disease-annotation of miRwalk for Hepatocellular carcinoma has been implemented for search, as combinatorial targets (supplementary 1). This analysis has evolved a list of HCC genes, further directed to determining the expected Cytoscape networking with our 24 mirs. Interestingly, the regulatory network of 24 mirs with its target genes has been overviewed in (figure 4).

**Figure4:**
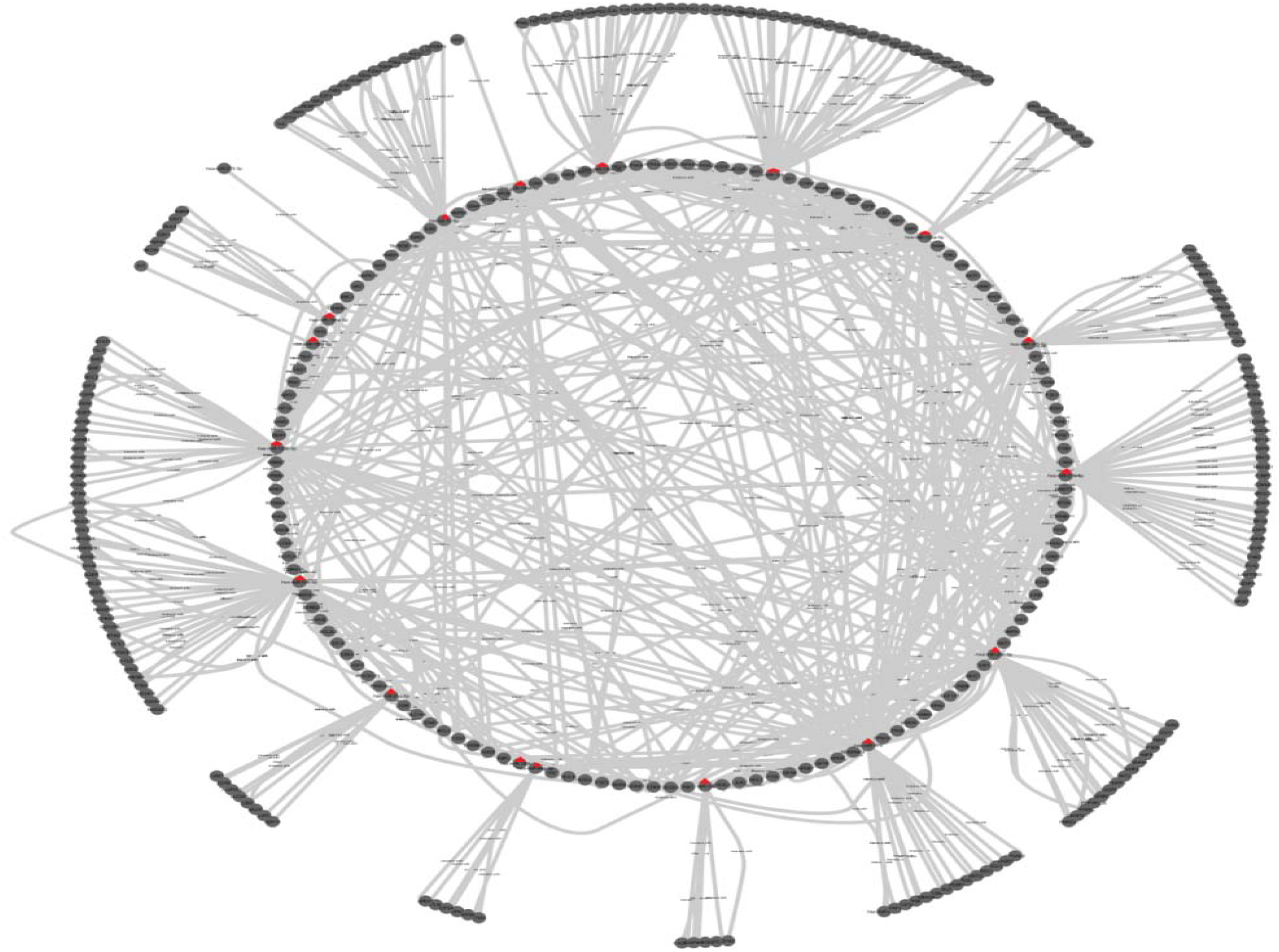
A regulatory network of microRNAs, Red triangle, with their targets, drawn by Cytoscape.

### 3.4. Biological Process of microRNAs

It was unveiled that microRNA has a biological process related to tumor suppressor and tumor activator (Figure 5).

**Figure5:**
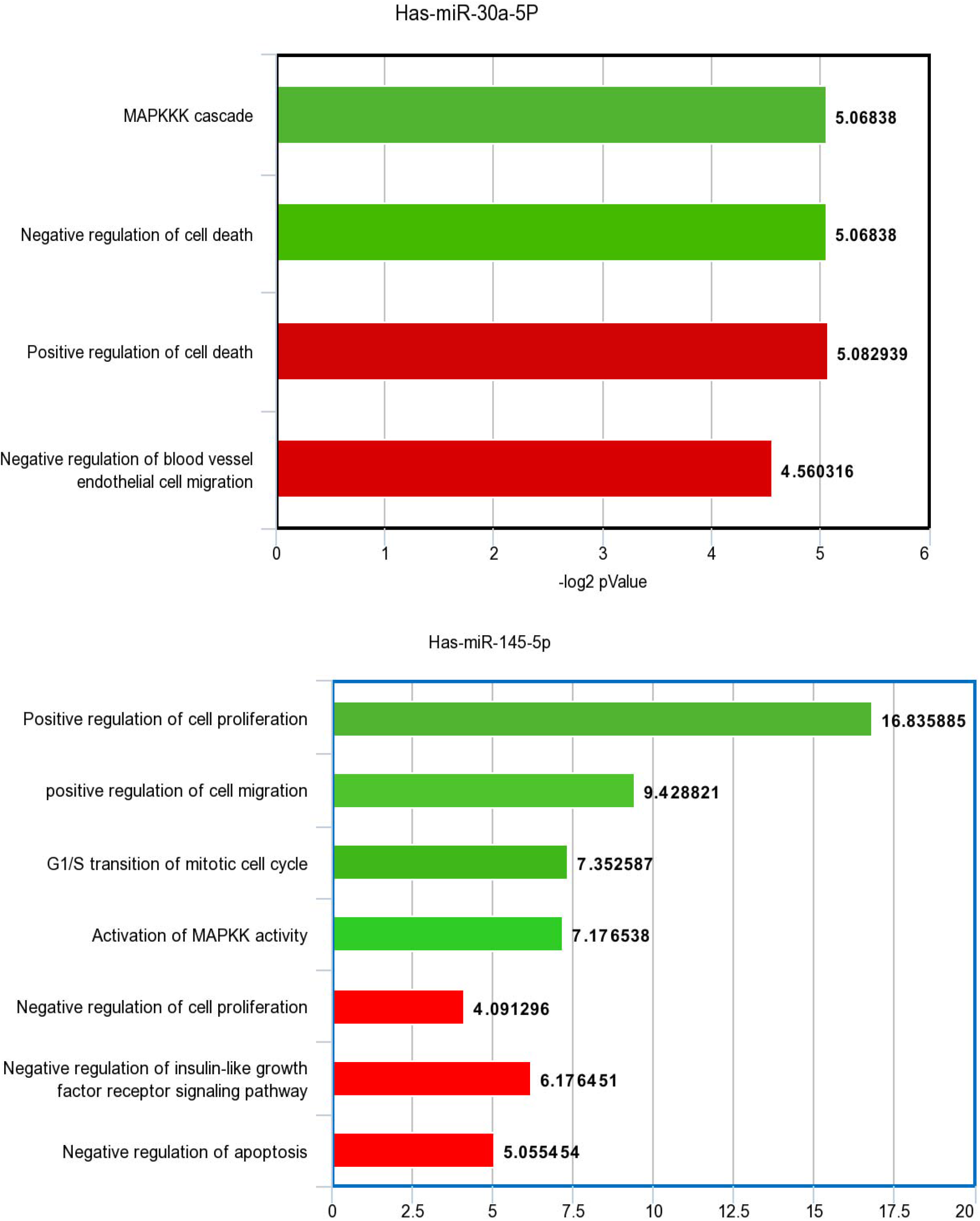
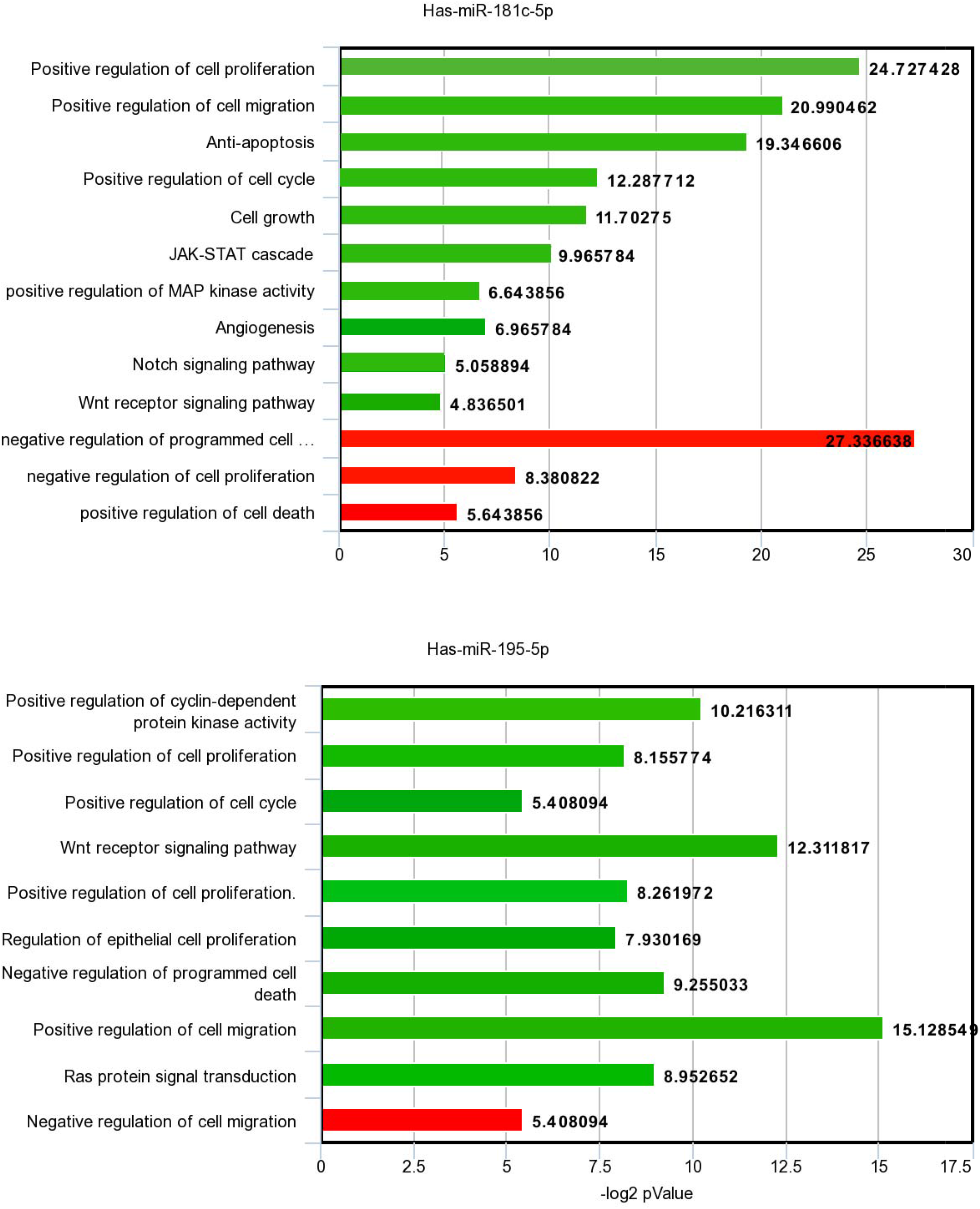
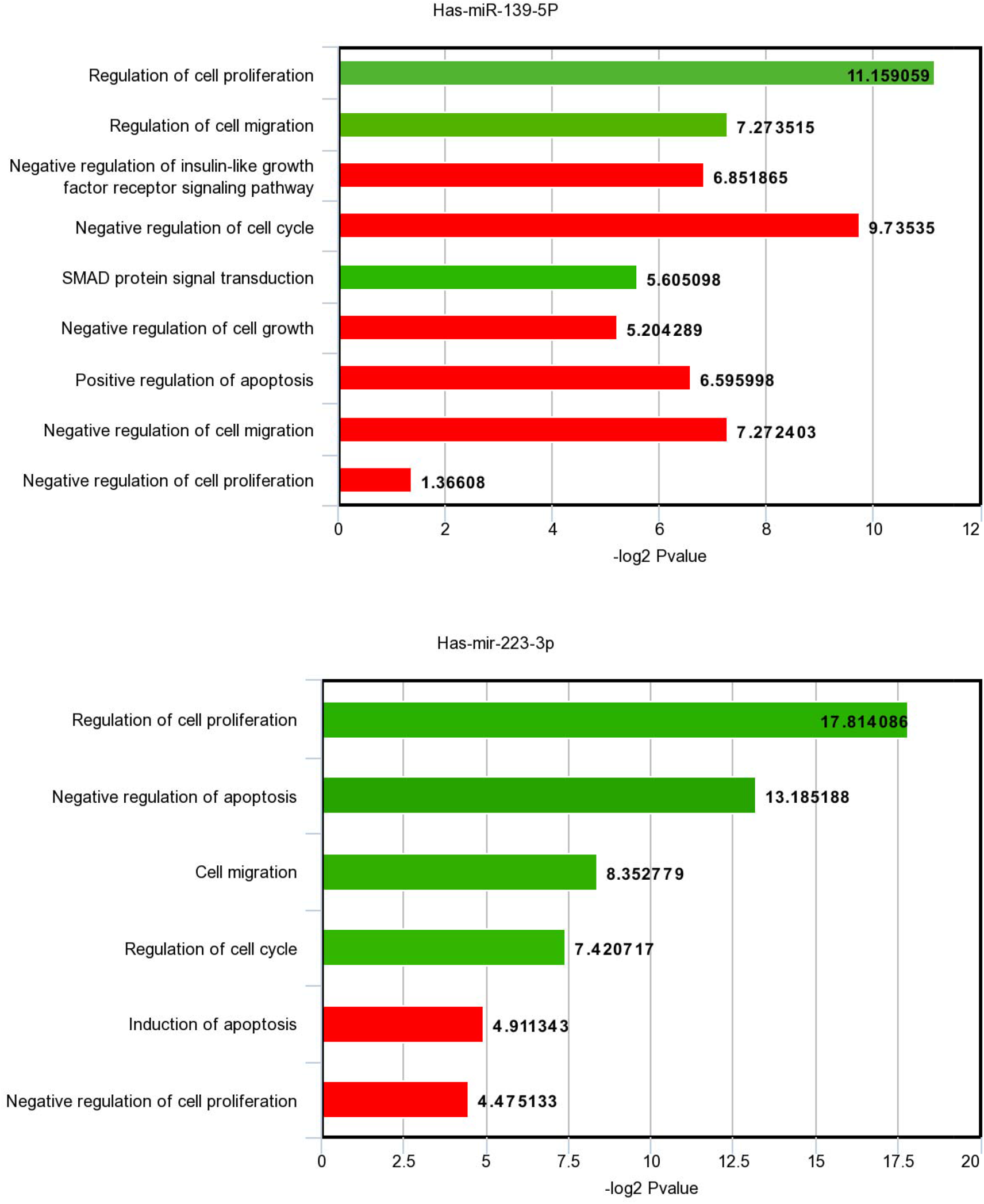
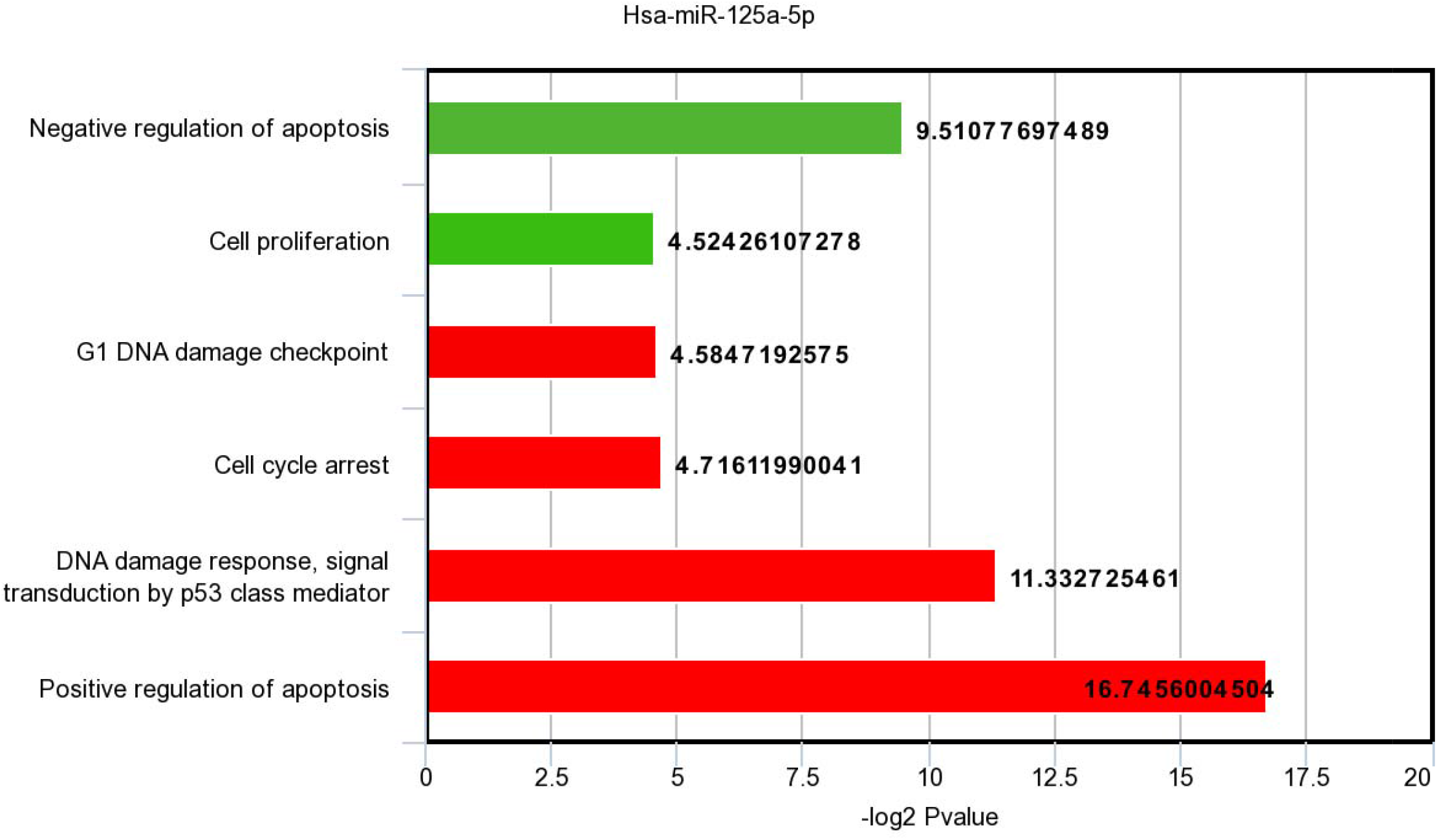
BP of target genes of down expressed miRs in HCC. Green bars show the tumor suppressor activity of miRs. Red bars show their oncogenic activity.

Here, figure 4 shows a set of down-expressed microRNAs in HCC with their biological process of the genes targeted by microRNA; only the BP of *P*-value < 0.05 has been selected. The dual role of microRNA in HCC is evaluated by the impact of microRNA on BP of HCC hallmarks either as an inducer or suppressor. The resultant is that each microRNA shows a dual role, either tumor suppressor (green bars) or onco-miR (red bars).

### 3.5. miRNA-TFs regulatory role

Four microRNAs have been found with abundant TFs targets (Figure 6).

**The figure 6:**
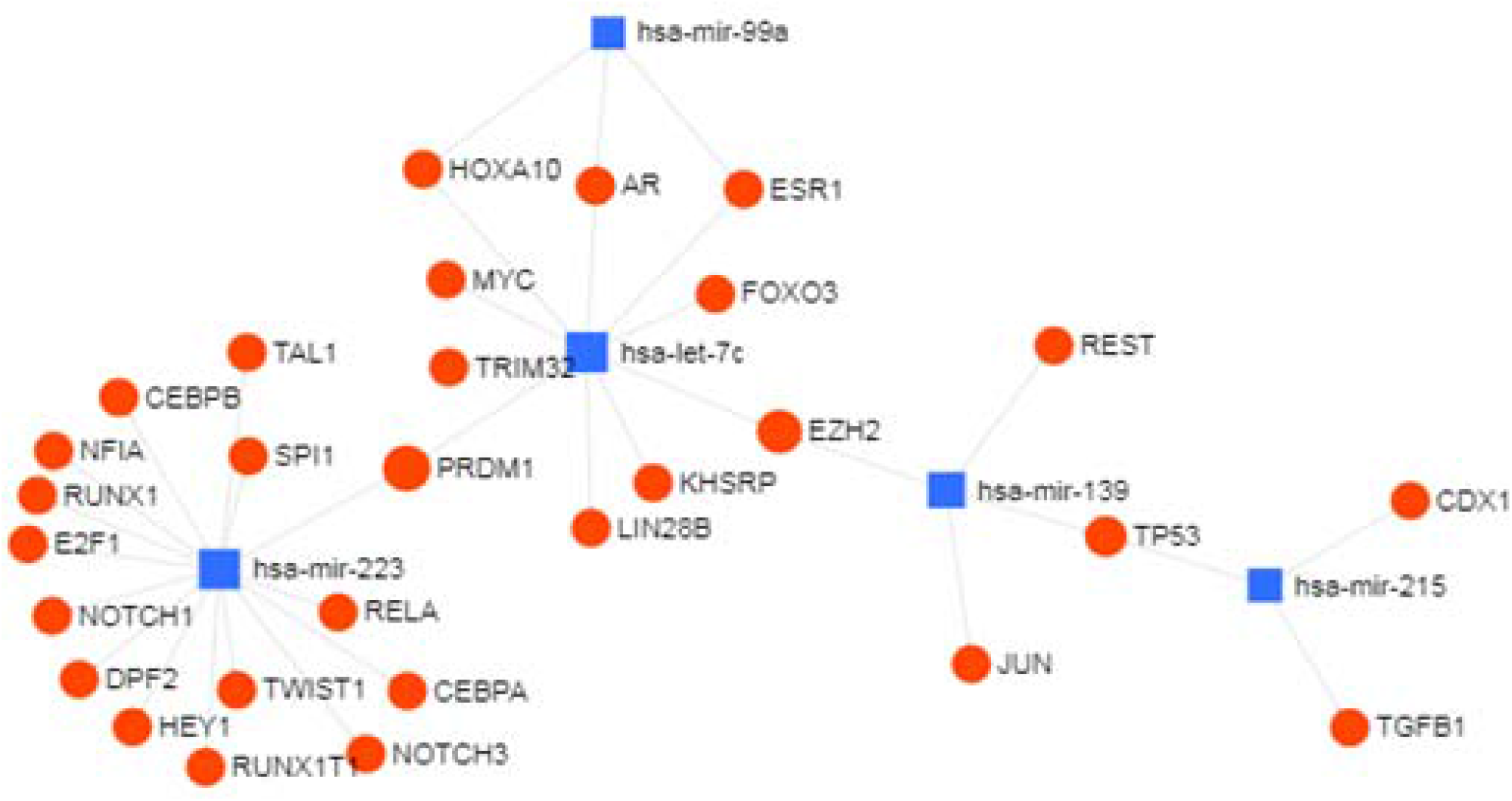
miRNA-TFs regulatory role.

Four microRNAs show tumor suppressor activity upon targeting TFs as transcriptional regulatory factors as ant-apoptosis and proliferation regulation. Hence, repressing these TFs results in tumor suppression, as shown in (table 1).

**Table 1:** shows Regulatory role of miRNA-TF interaction in HCC

| MiRNA | TFs | Regulatory role of miRNA-TFs interaction in HCC |
| --- | --- | --- |
| hsa-let-7c | AR,ESR1,EZH2,FOXO3,HOXA10,KHSR<br>P,LIN28B,MYC,PRDM1,TRIM32 | Tumor supressor |
| hsa-mir-139 | EZH2,JUN,REST,TP53 | Tumor supressor |
| hsa-mir-215 | CDX1,TGFB1,TP53 | Tumor supressor |
| hsa-mir-223 | CEBPB,DPF2,E2F1,HEY1,NFIA,NOTCH<br>1,NOTCH3<br>,PRDM1,RELA,RUNX1,RUNX1T1,SPI1,<br>TAL1,TWIST1 | Tumor supressor |

### 3.6. miRNA-LncRNA interaction

Eight microRNAs have shown potential interaction with lncRNAs (Figure 7) (Table 2). LncRNA acts as a CeRNA sponge for microRNA. Subsequently, it results in the excessive inhibitory activity of miRNA-mRNA interaction by excessive down-expressing of miRNAs.

**Figure 7:**
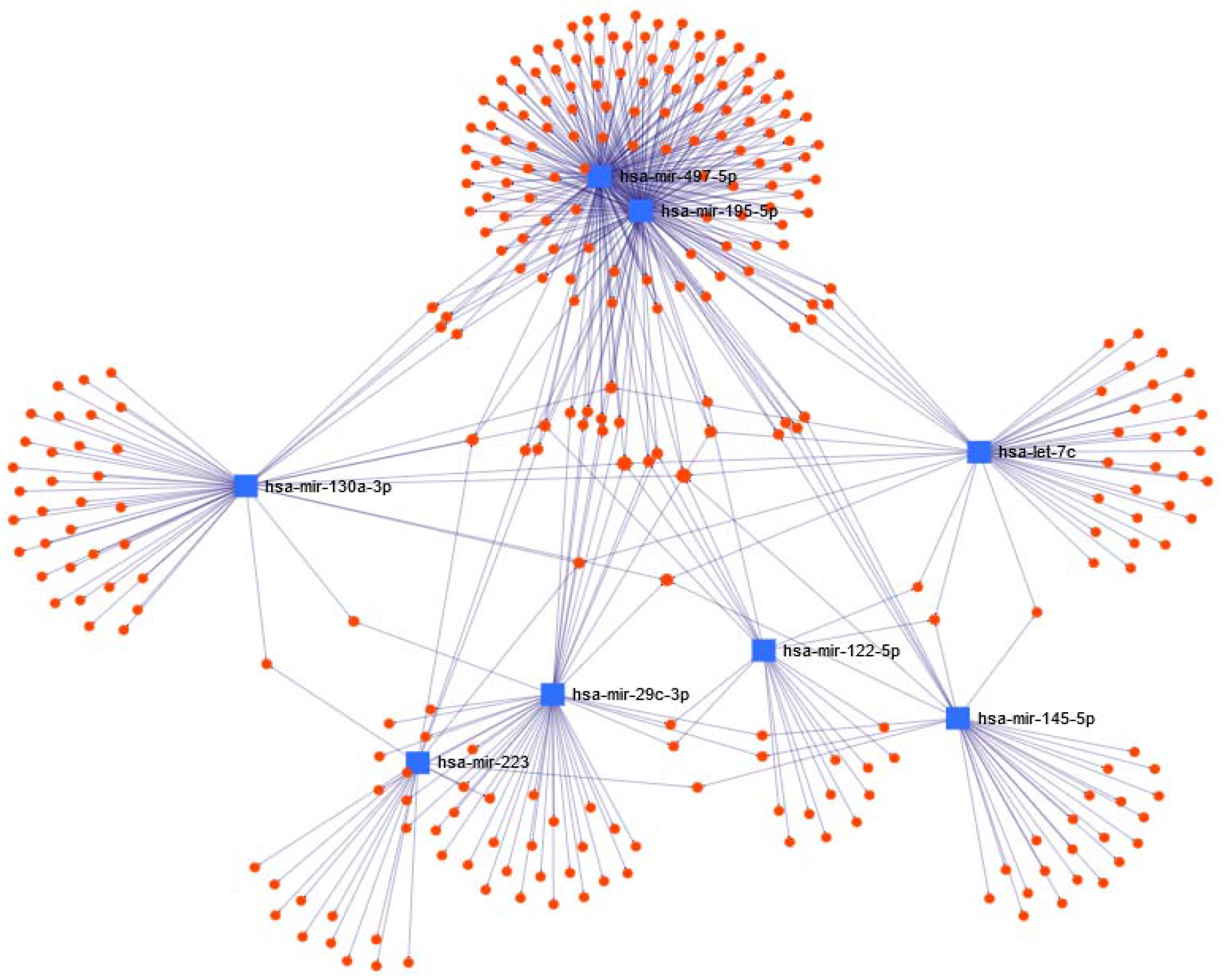
miRNA-lncRNA regulatory role.

**Table 2:**
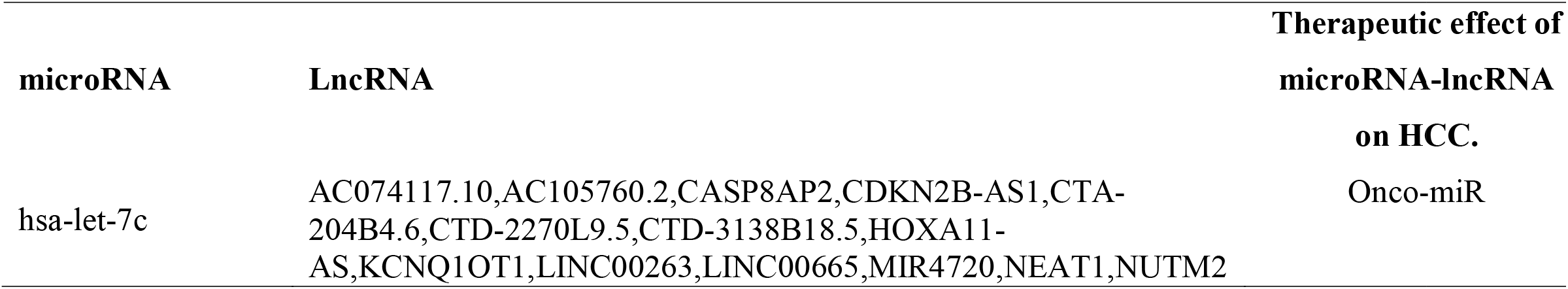
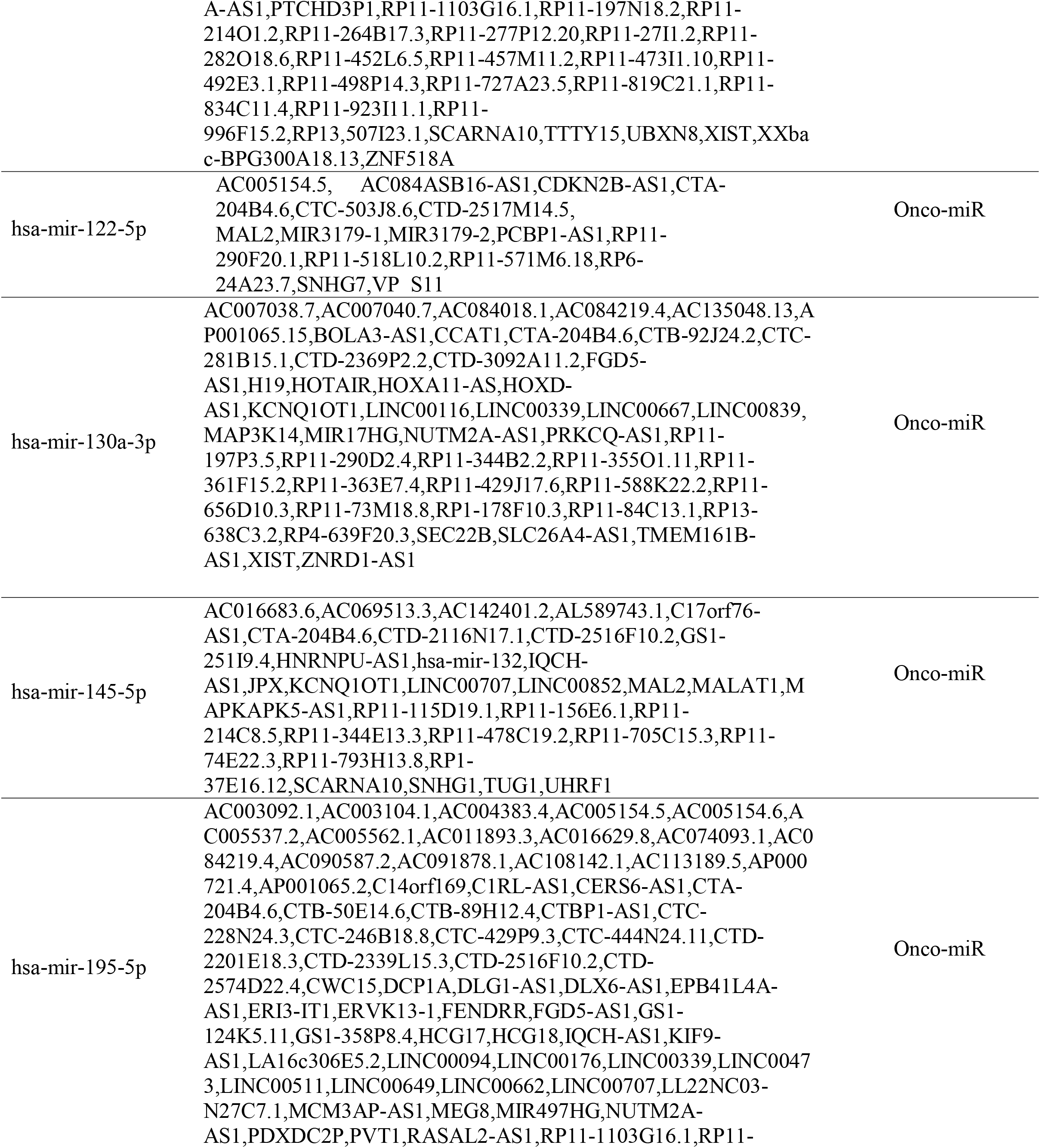
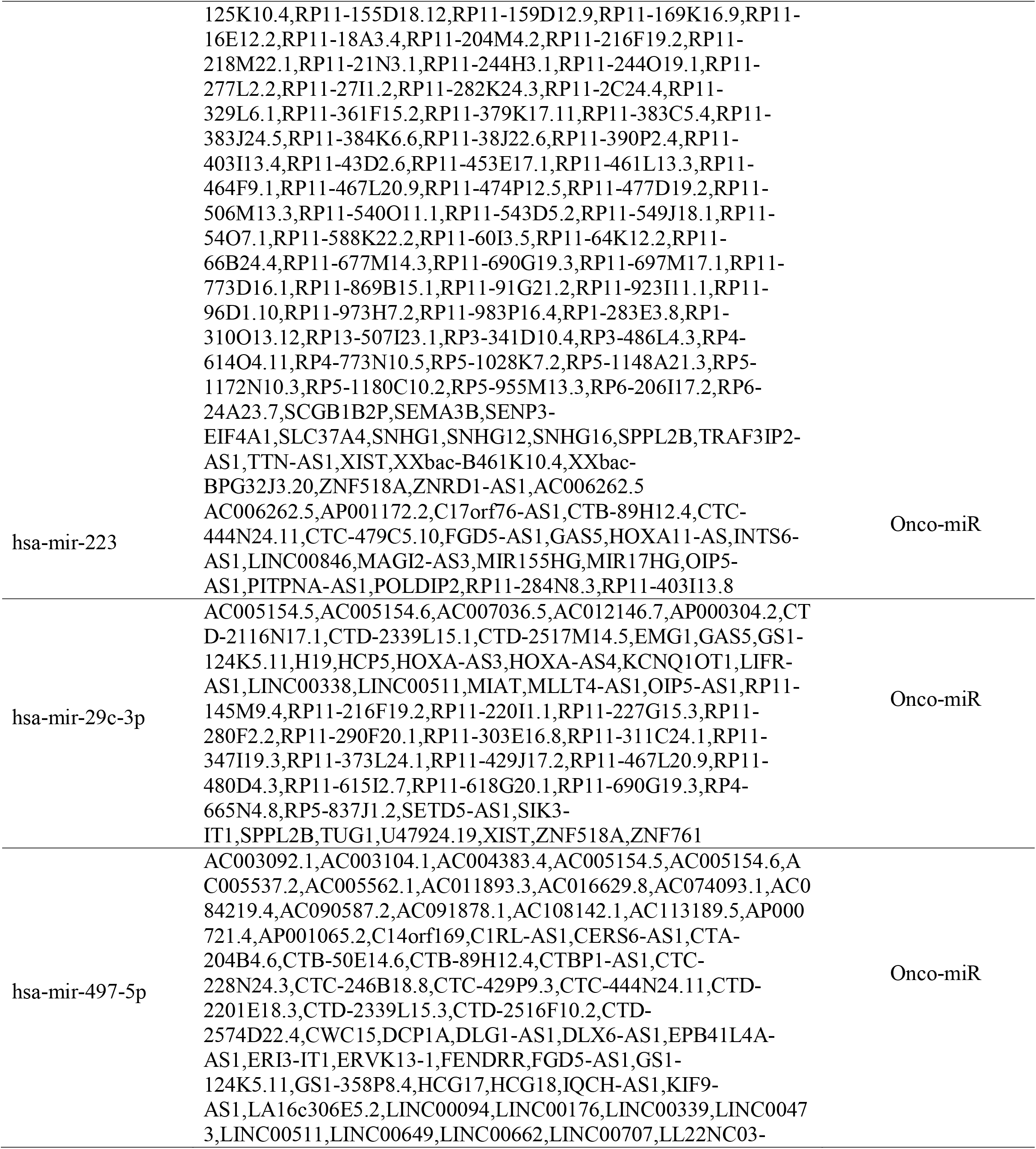
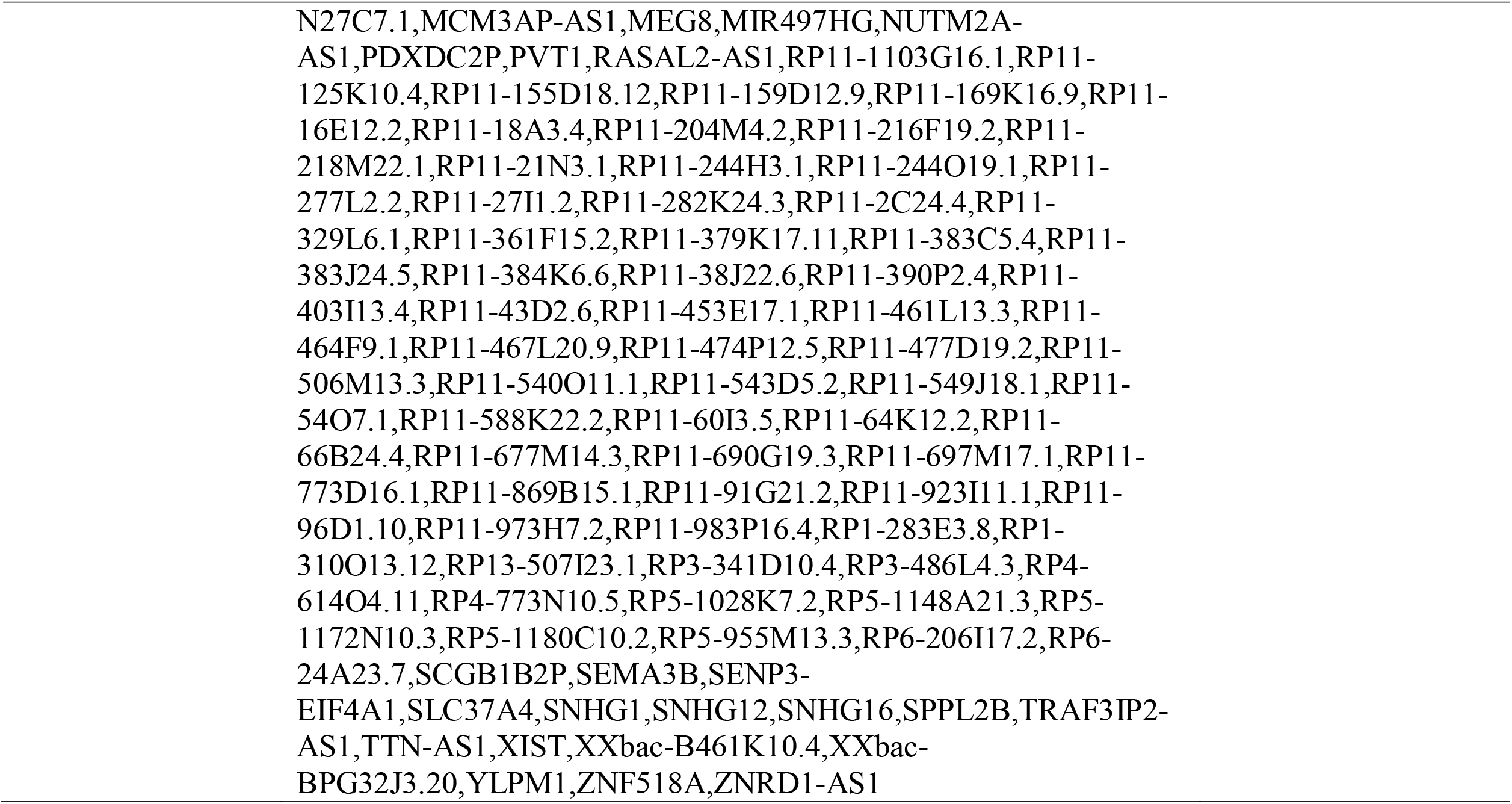
shows Regulatory role of Therapeutic effect of microRNA-lncRNA on HCC

### 3.7. miRNA-miRNA interaction

Here, it is predicted shown that microRNA are acting as ceRNA for each other’s (Figure 8).

**Figure 8:**
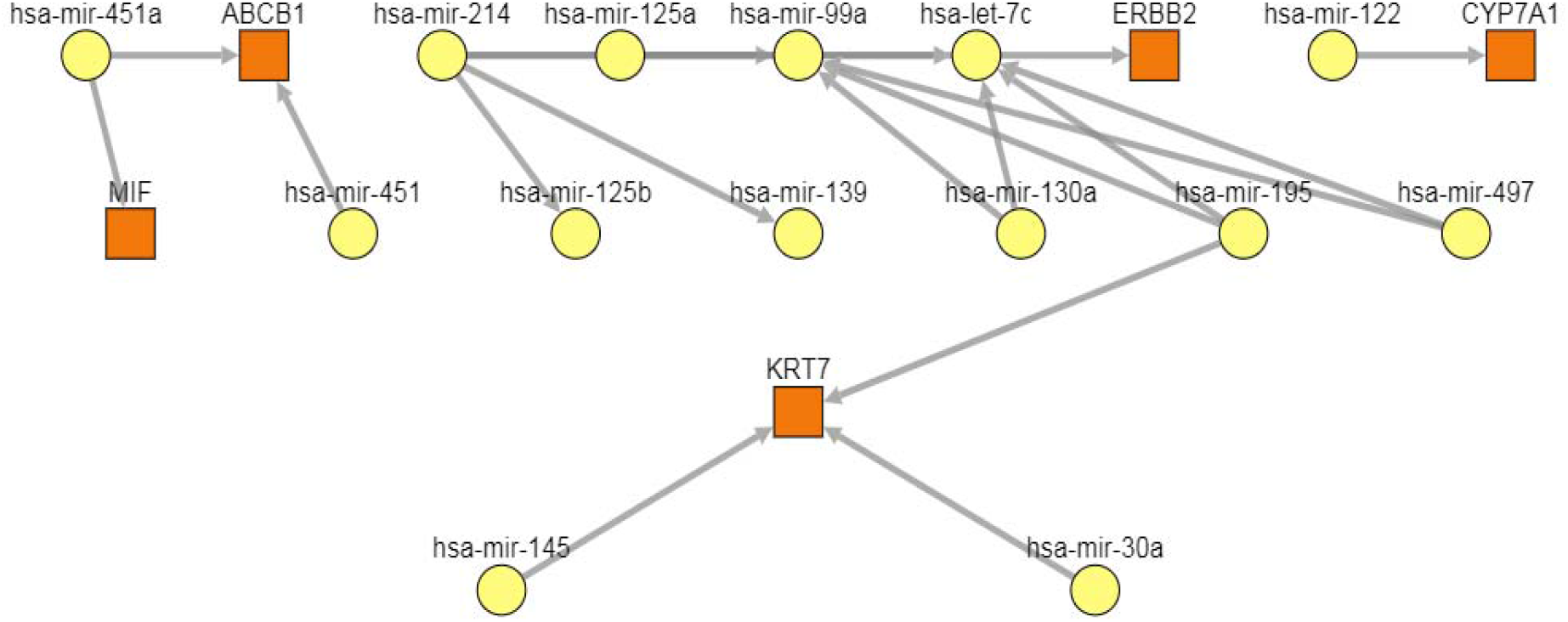
miRNA-miRNA, PmmR, regulatory interactions with 10 nodes and 19 edges. Here, all the input microRNAs are supposed to be therapeutically tumor suppressor. However, these microRNAs acts as a sponge to each other. Consequently, it is putative that this kind of interaction results in an oncogenic impact.

It results in enhancing microRNA regulatory role as tumor suppressor (Fig.7). Seven miRs show ceRNAs activity.miR-214 acts as sponge for (miR-125a, miR-99a, miR-139, miR-125b).miR -125a acts as sponge for miR-99a.miR-99a acts as sponge for let -7c.miR-130a acts as sponge for (let-7c, mir-99a).miR 195 acts as sponge for (let-7c, miR-99a).miR-497 acts as sponge for (let-7c, miR-99a).

### 3.8. microRNA-PPI interaction

Six major biological processes in HCC were resultants (Figure 9). These BP are GO:0051726 regulation of cell cycle, GO:0071158 positive regulation of cell cycle arrest, GO:1901991 negative regulation of mitotic cell cycle phase transition, GO:0040008 regulation of growth, GO:0042981 regulation of the apoptotic process, GO:0043066: Negative regulation of the apoptotic process. As shown in table 3, GO:0051726 (regulation of cell cycle) BP has 30 genes as targets for ten down expressed miRNAs in HCC. Because activation of the cell cycle is one of the major hallmarks of liver cancer. Hence, suppressing the cell cycle by miRNAs results in considering that activity as miRNA tumor suppressor. On the other hand, GO:0071158 (positive regulation of cell cycle arrest), GO:1901991 (negative regulation of mitotic cell cycle phase transition), GO:0042981 (regulation of apoptotic process) BPs have 7, 15, 27 genes, respectively, which are non-incorporated in HCC. It is because GO:0071158, GO:1901991, and GO:0042981 BPs are not hallmarks of causing liver cancer. Therefore, suppression of GO:0071158 by miRNAs results in oncogenic aspects.

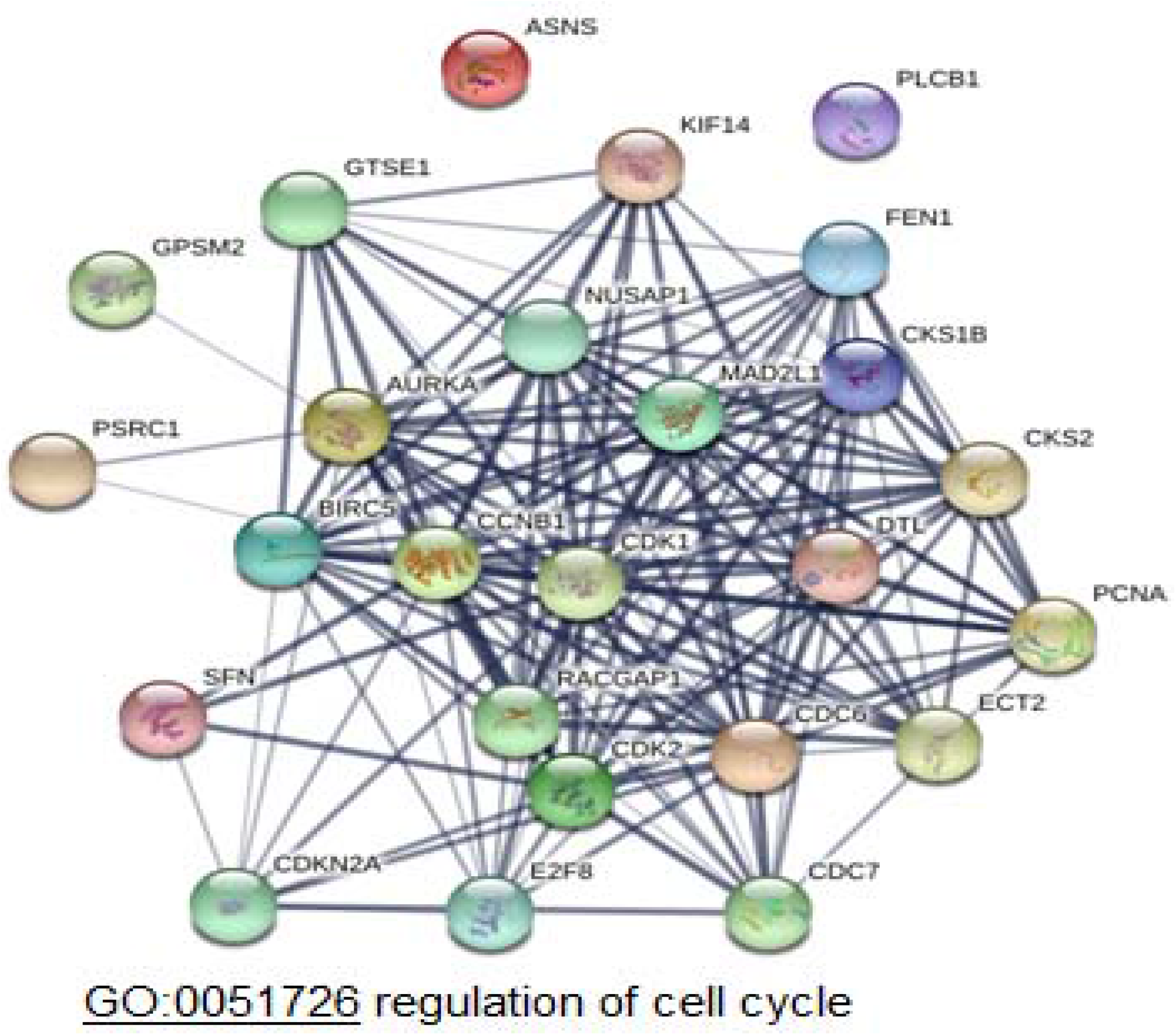
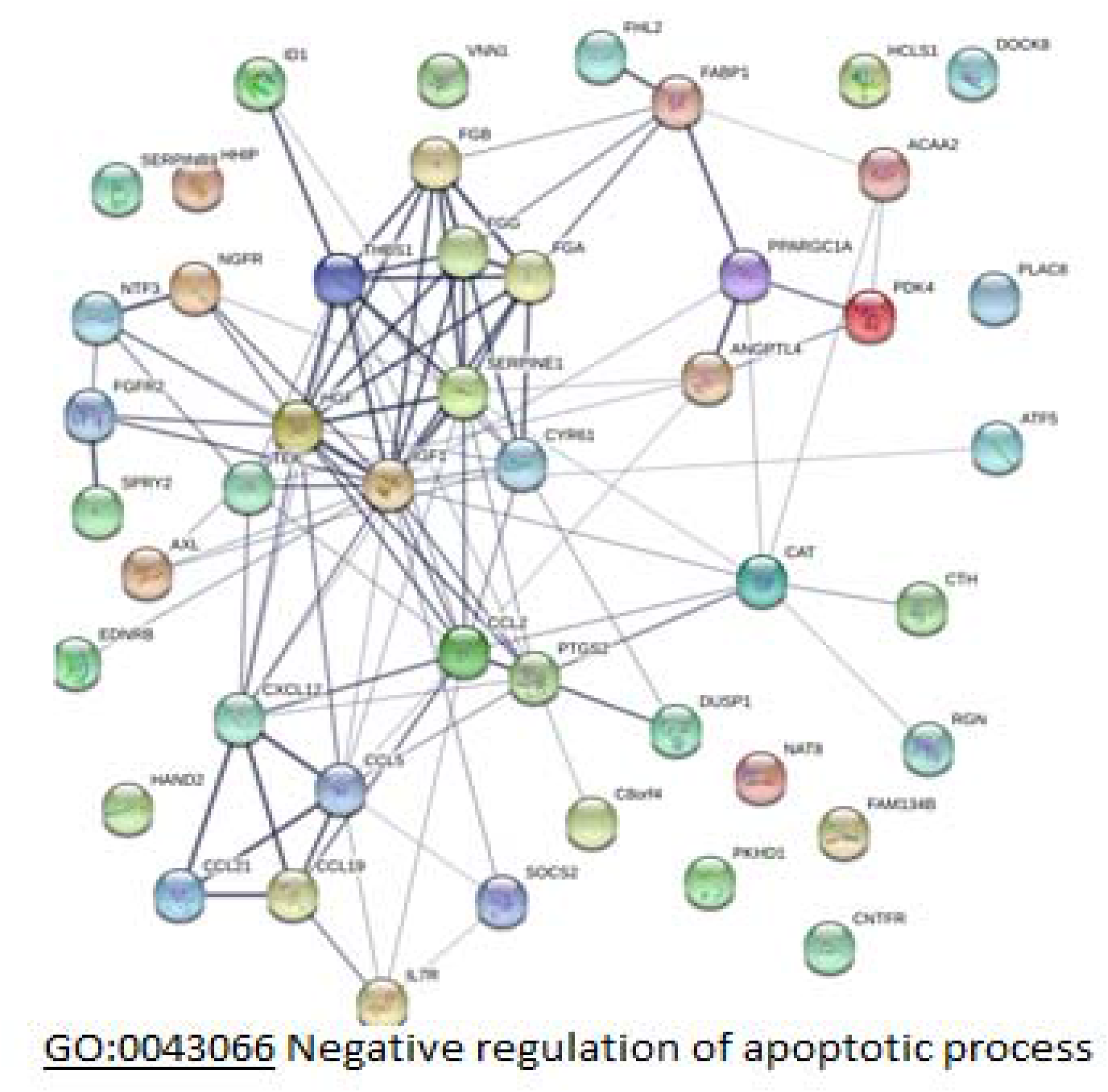
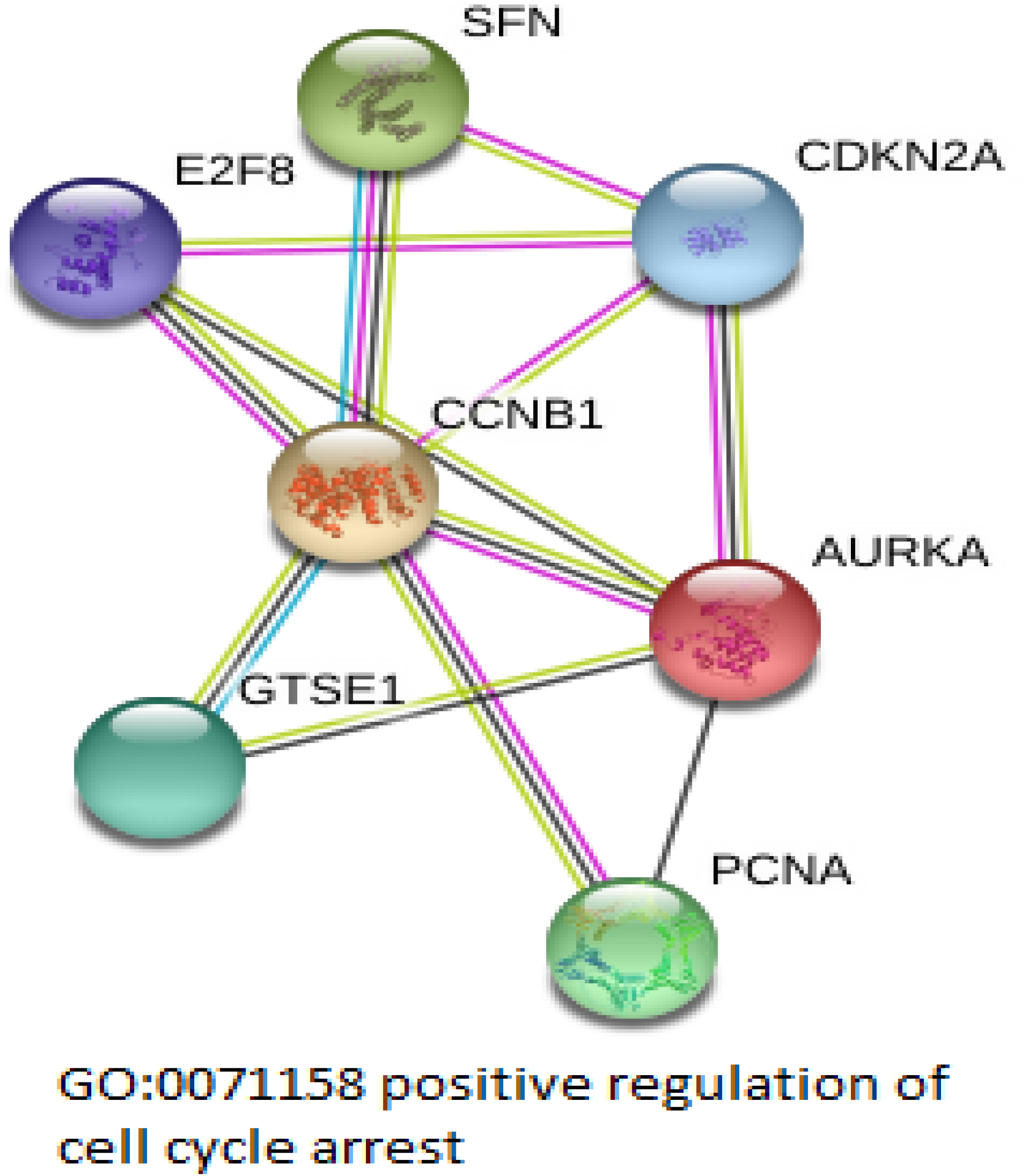
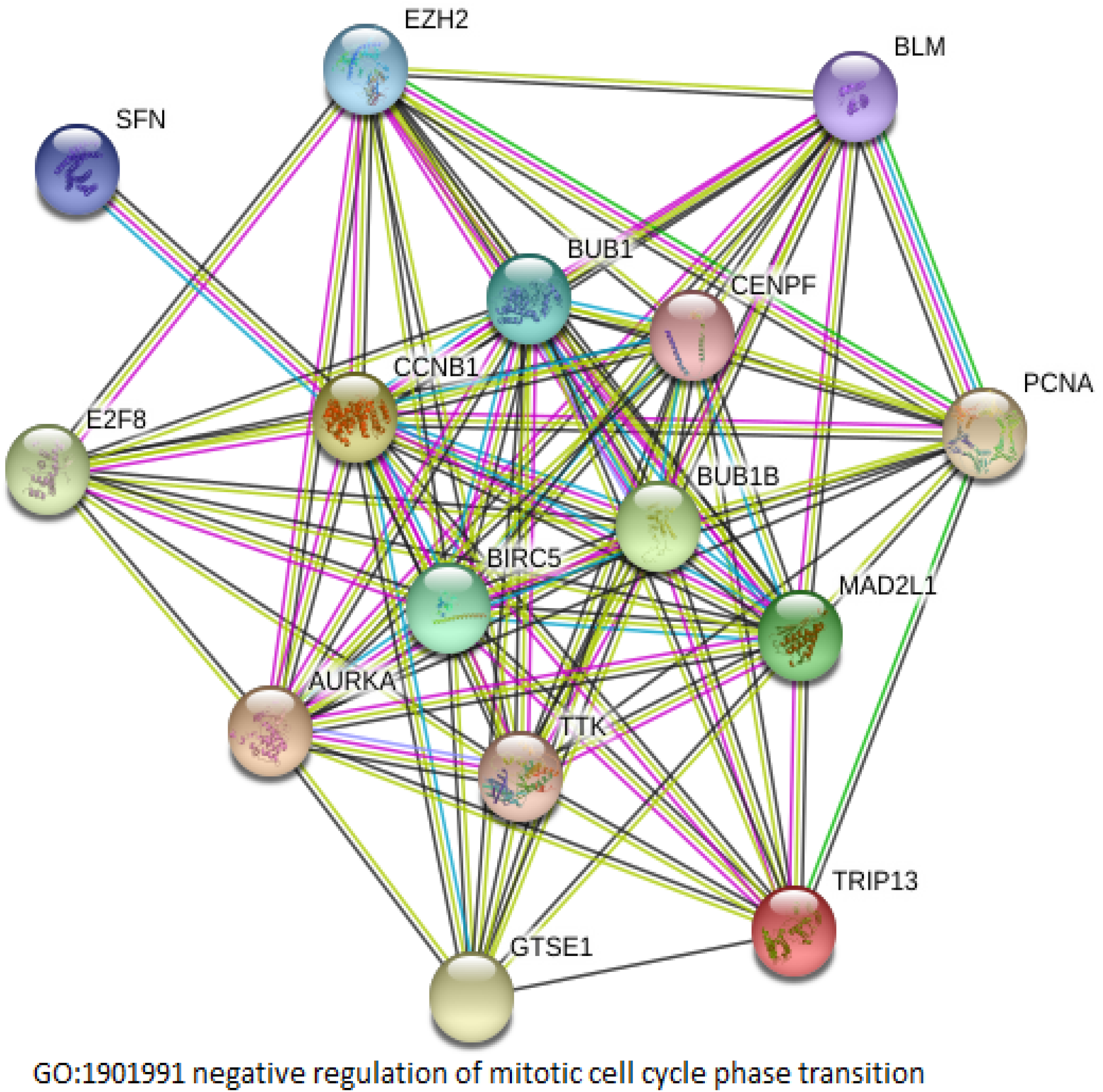
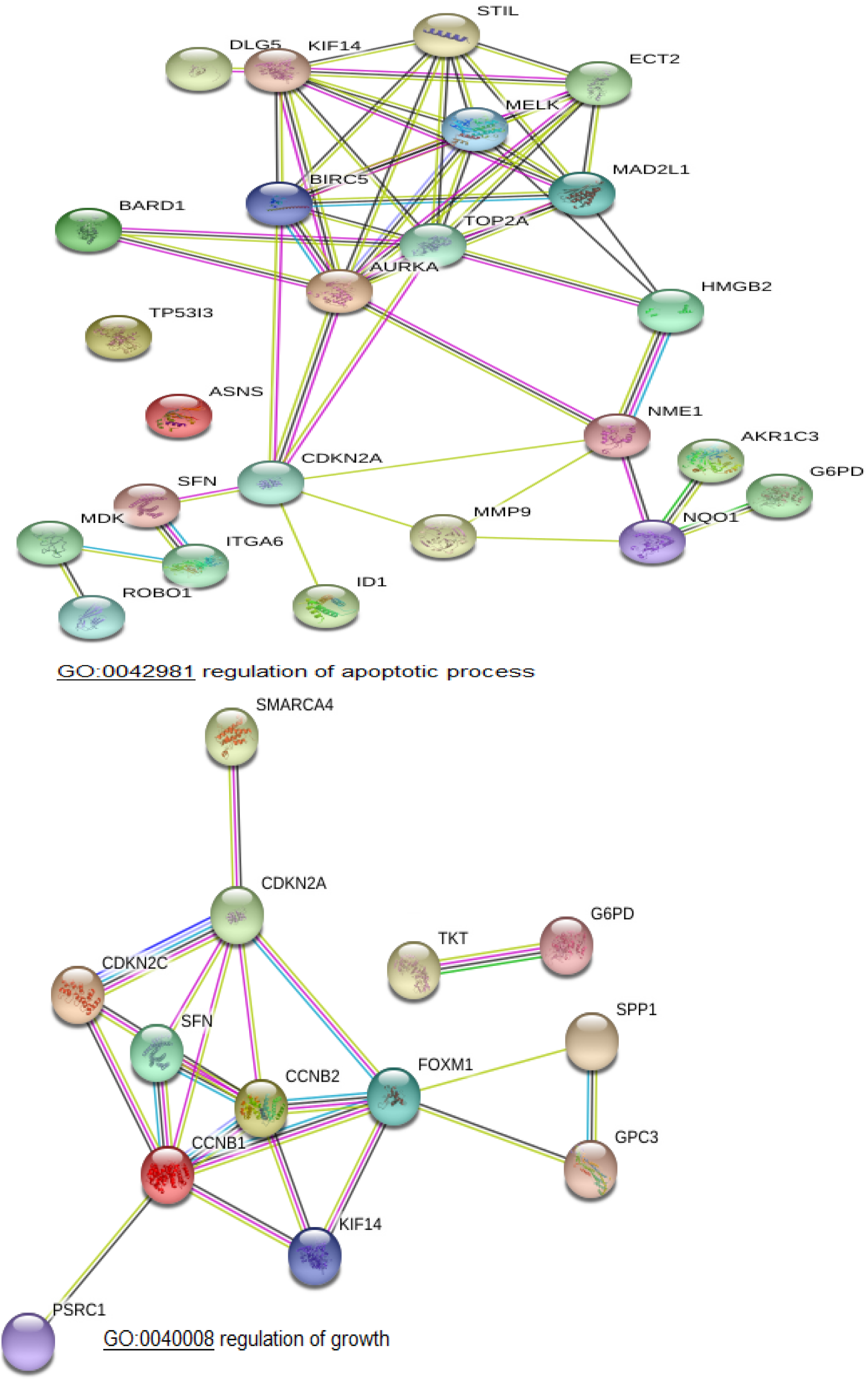

**Table 3:**
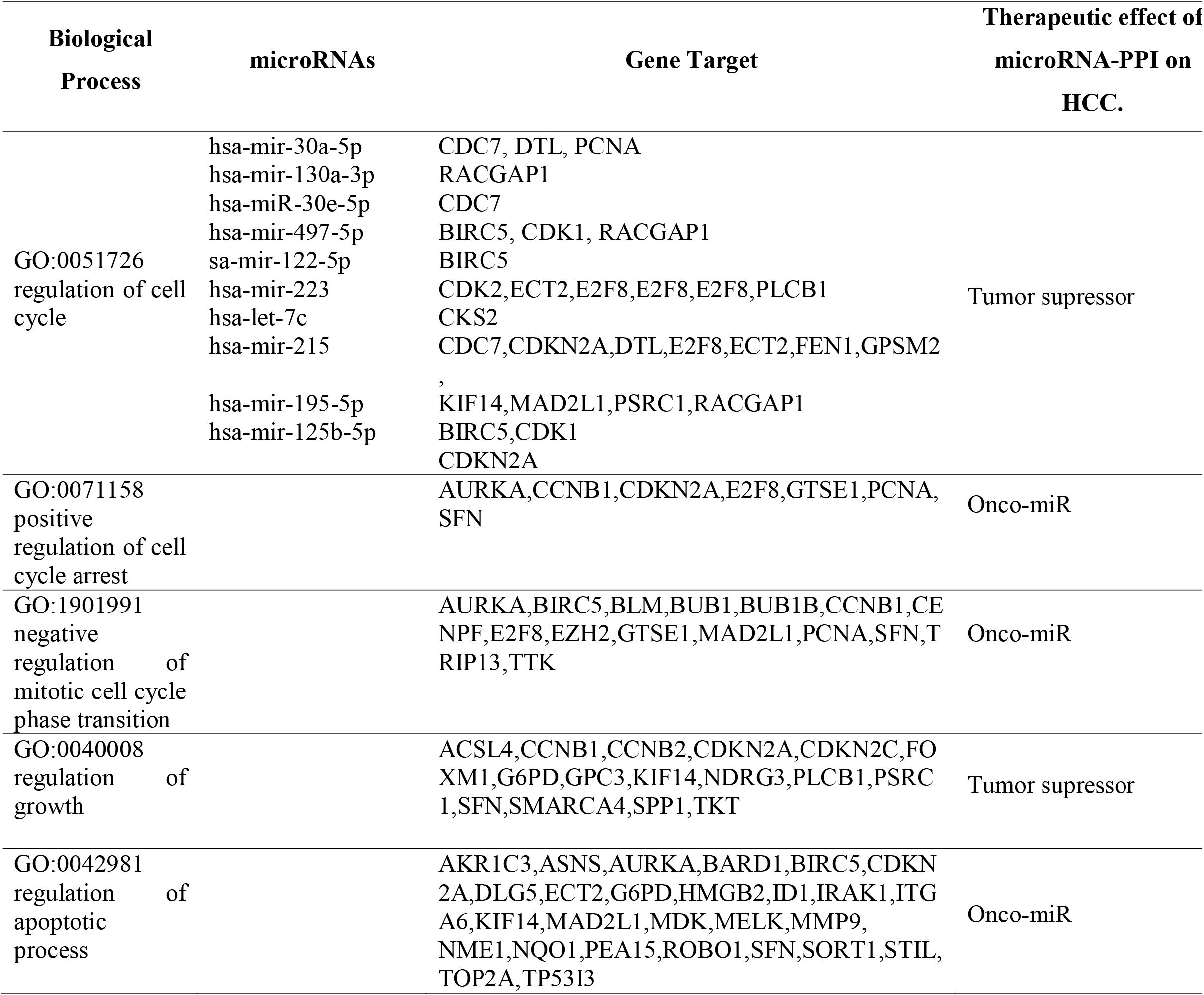
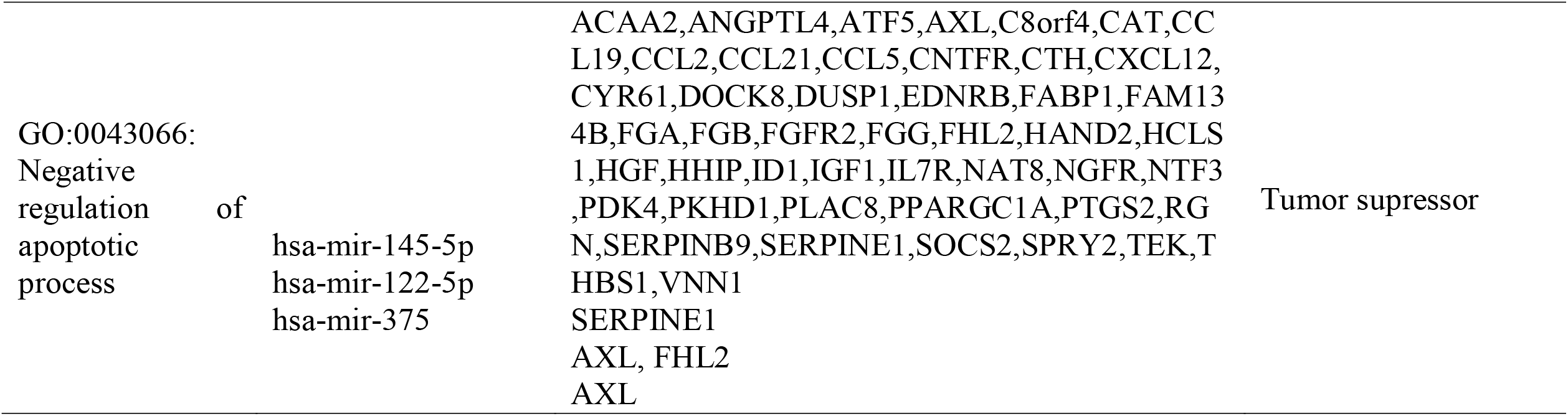
Illustration of the Biological process of PPI network and their matched microRNAs

Figure 9 shows biological processes retrieved from PPI network of 180 overexpressed genes and 572 down expressed genes in HCC.PPI network was analyzed by STRING.

### 3.9. Free survival analysis

Here, survival analysis shows us that ectopic expression of (CCNB1, AURKA, BIRC5, HGF, IGF1, TOP2A) has poor survival progression in the long term in LIHC(Figure 10). It means that microRNA acting on these genes is non-additive value for curing HCC in the long term.

**Figure 10:**
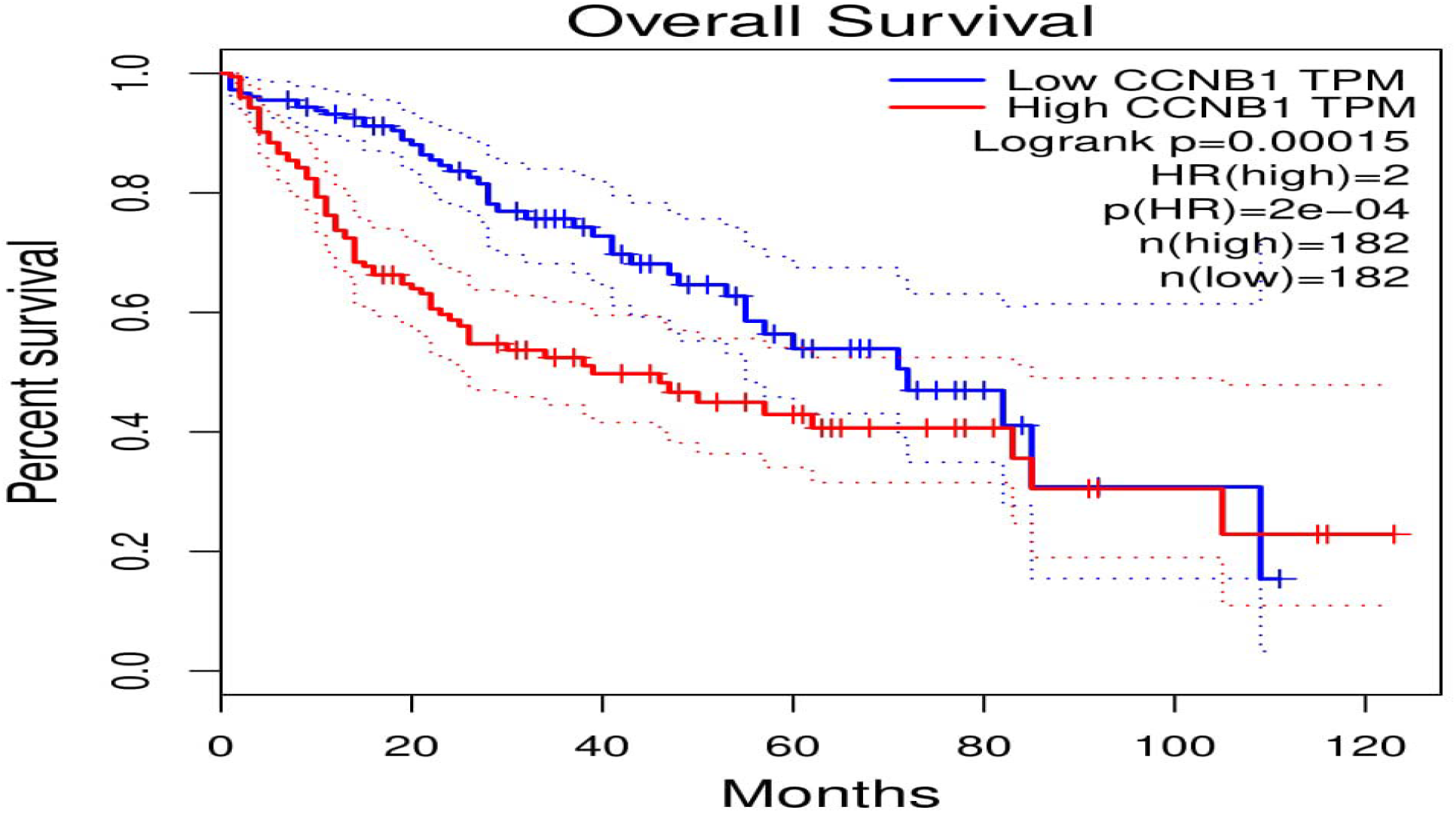
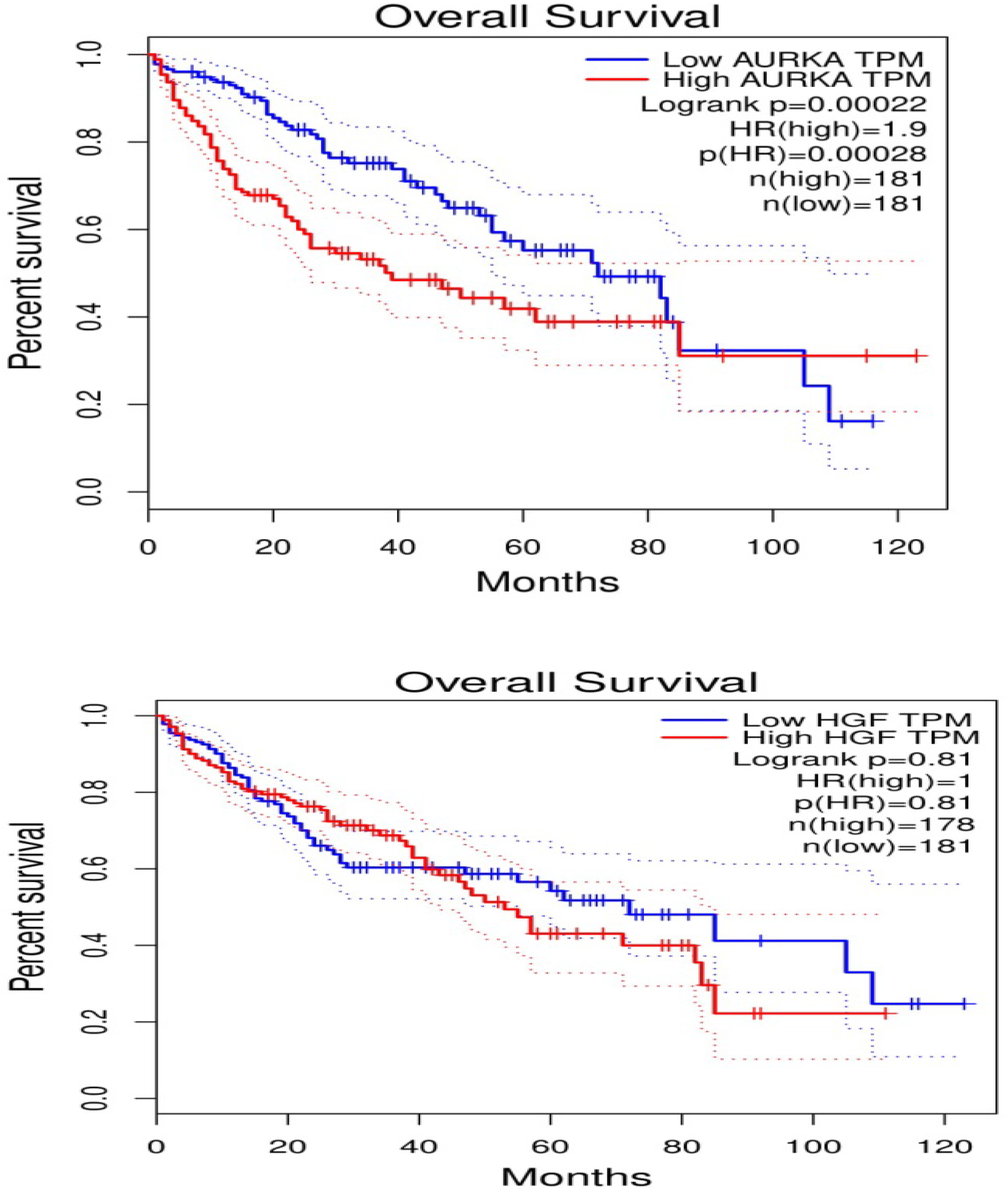
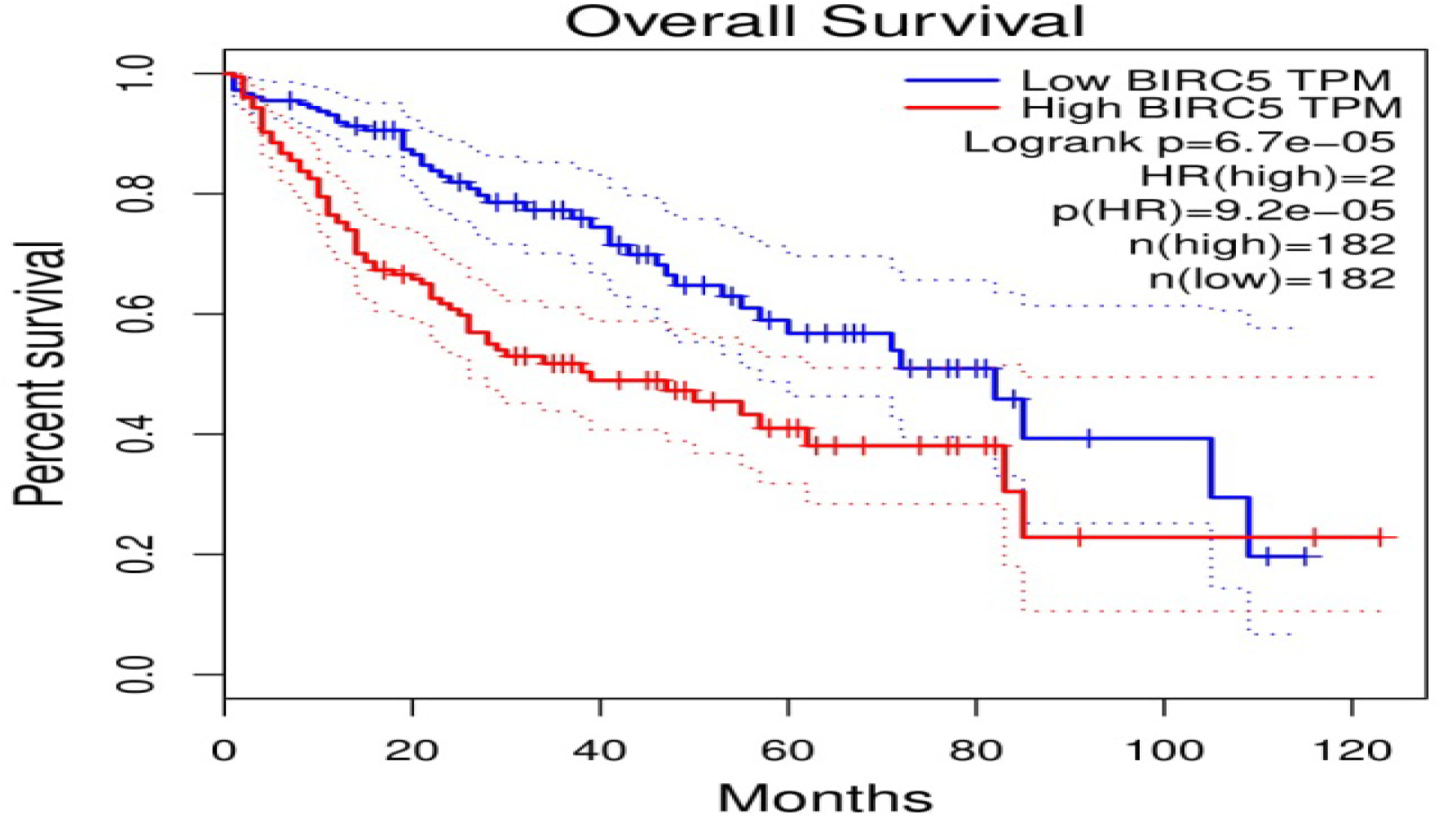
list of survival figures of the gene target.

## 4. Discussion

MicroRNAs are a promising therapeutic gene therapy for Cancer (Table 4).

**Table 4:**
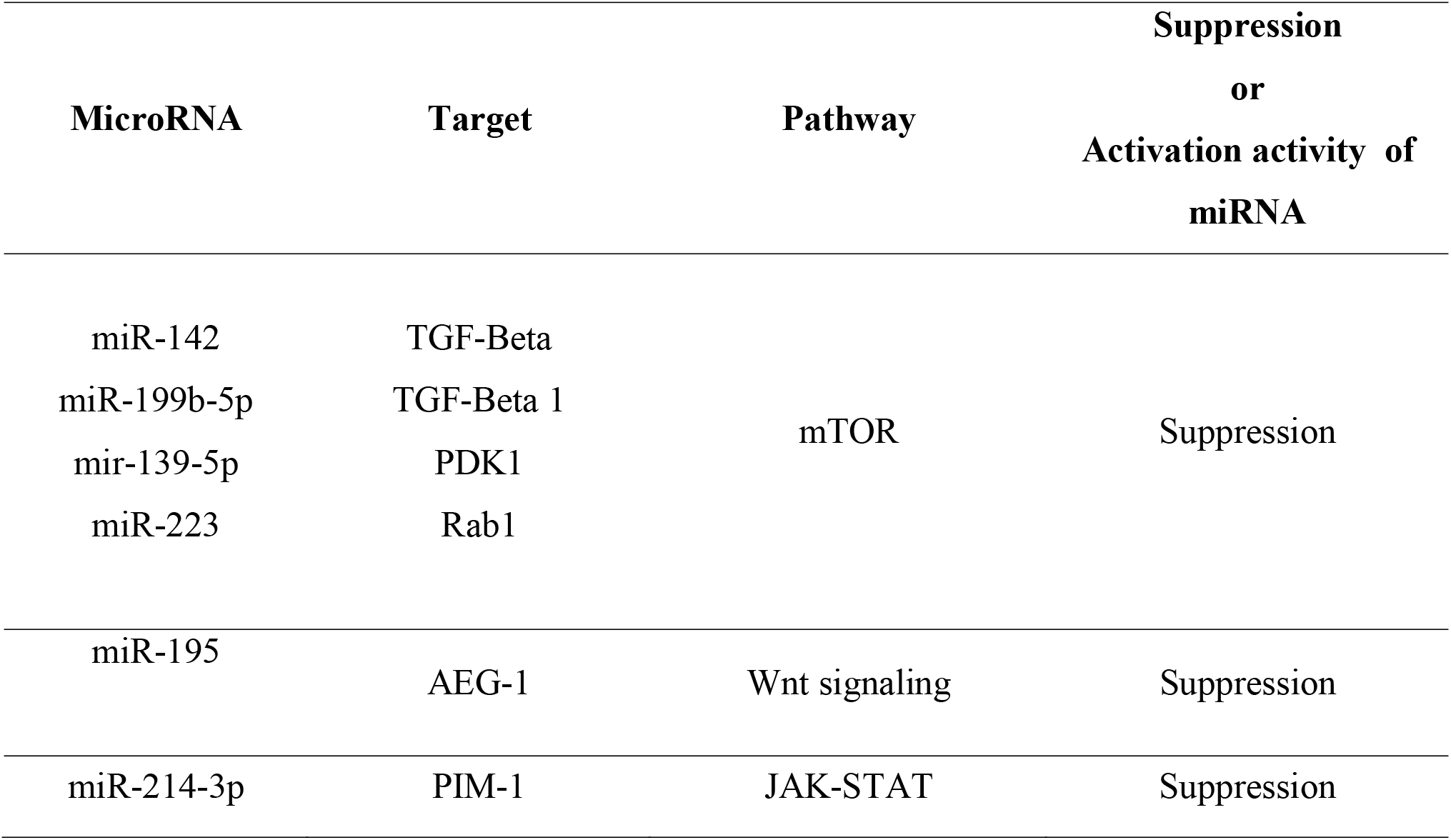
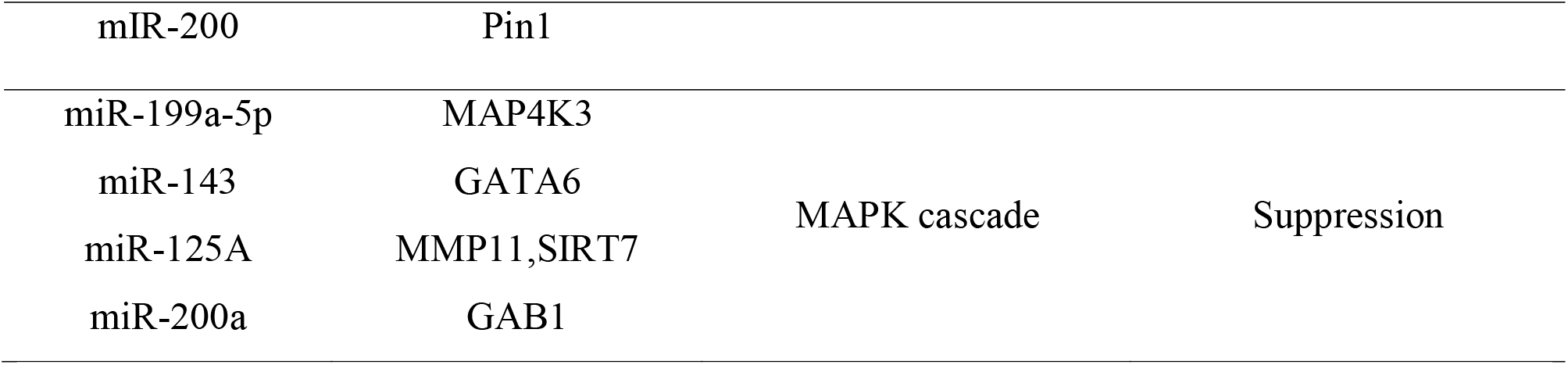
list of miRNAs as HCC therapy.

They are involved in regulating many biological processes. Their ability to bind with 3’ untranslated region of mRNAs recruit many factors to degrade mRNAs, consequently acting as a repressor for oncoprotein synthesis [48, 49]. It results in knocking down the pathologically causative genes of Cancer [50] (Table 3). However, they have putatively mysterious functions of acting through inversely dual role either as a tumor suppressor or oncomiR. Through our current work, we tried to uncover the dual role of a set of microRNAs in HCC. MicroRNAs, on the contrary to siRNAs, lack specificity to bind mRNAs, where MicroRNAs have binding seed regions with a high capacity to target multiple genes [51]. It is considered a drawback in microRNAs as an approach to gene therapy. It is experimentally invalid to target specific oncogenes to be knocked down by microRNA where microRNAs target tumor suppressor genes, off-targets, that result in activating cancerous biological processes. To uncover these dually mysterious microRNA functions, we have collected the common down-expressed genes of miRNAs HCC within different GEO and TCGA-LIHC databases. The chosen databases are for (HCC vs. cirrhotic non-cancer) and (G2/M phase of the cell cycle in vs. normal tissues) to cover different pathological stages of HCC.

Additionally, these datasets cover broad samples (446 HCC vs. 146 normal specimens).In this respect, we targeted only the microRNAs common in all the chosen datasets due to the inconsistency of the results as miRNA measurement technology is immature compared to gene expression’s system where CapitalBio’s is the least reliable dataset. Milteny’s and TCGA’s datasets generated the most consistent results. Affymetrix’s arrays are weak in detecting the expression difference. Hence, we decided to integrate these datasets to get the common down-expressed miRNAs to ensure accuracy.

Our work aims to support the combinatorial therapy of microRNA. One technique activates the tumor suppressor genes, off-target genes, via designing a cassette of miRNA-guide RNA-CRISPR/Cas9 [52]. Maybe CRISPRa is a choice to be combined with microRNA to activate these off-targets genes knocked down by microRNAs. An additional suggestion is to combine microRNA with an exogenous expression vector carrying the tumor suppressor genes. Also, combined tumor suppressor microRNA with those microRNAs that activate tumor suppressor genes via binding its 5’UTR is a third way to solve the drawback of microRNAs’ dual role [53]. The idea behind our suggested combinatorial therapy is that it helps most in increasing the level of expressed tumor suppressor genes within the cells.

Moreover; microRNA cocktail therapy should not be a combination of microRNAs which act as CeRNA for each other [54]. One example of the dual role of microRNAs is “how microRNAs regulate dually in colorectal cancer?”; It was reported that miRNAs show dual regulatory roles in colorectal Cancer [55]. In a wet lab, much microRNAs act dually. miR-375 shows dual functions in prostate cancer [56].In Breast cancer, miR-181a acts as an onco-miR via targeting Bax, increasing metastasis. On the other hand, it acts as a tumor suppressor via targeting BCL-increasing apoptosis [57].

The final regulatory role of microRNA is evaluated by miRNA-LncRNA-mRNA-TFs Crosstalk as an accumulative impact on how microRNA acts (Figure 11).

**Figure 11:**
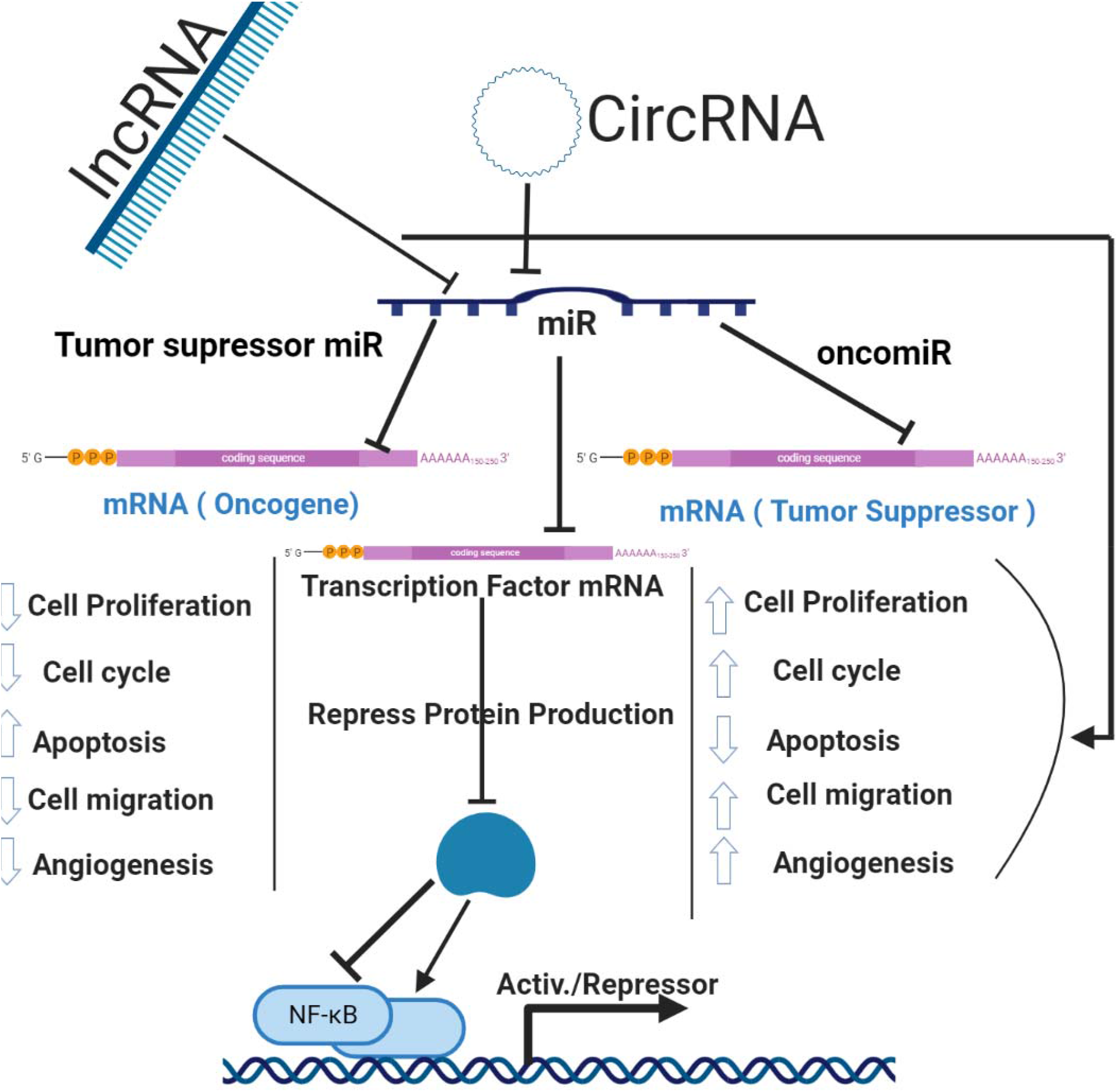
Regulatory role of microRNA as either oncomiR and/ or tumor suppressor miR.

LncRNA is 200 nt, acting as epigenetic regulators and as competing RNA for microRNA. It results in decreasing in the affinity of microRNA to be available for mRNA. Subsequently, it is predicted to alter microRNA regulatory role shifting it to be oncomiR for down expressed miRNAs in Cancer. On the other hand, transcription factors act via either activator or suppressor for the genes. So, microRNA acting on TFs results in either enhancing its tumor suppressor activity or shifting into being oncomiR. It is based on the ontology of the gene where TF targets.

Through our work, we concluded that microRNAs are incorporated in most hallmarks of hepatocellular carcinoma. However, these microRNAs’ dual activity made the consideration of the dominance of one of both functions a tough decision. Hence, we strongly suggest that more investigations on the factors that affect which microRNA function types will predominate should be carried out to solve their Myth. We suggest enhancing the protocols of using microRNAs as a gene therapy by combing activating techniques with miRs. These activating techniques are to induce the tumor suppressor genes targeted by microRNAs.

## Data Availability

All the data is available by requesting from the corresponding author.

## Authors’ Contributions

Mahmoud M. Tolba, Bangli Soliman and Mahmoud Elhefnawi designed this project. Mahmoud M. Tolba and HebaT’Allah Nasser conducted the reseach. Mahmoud M. Tolba, Abdul Jabbar edited the manuscript.Mahmoud M.tolba designed the figures.

